# Long-term gut colonization with ESBL-producing *Escherichia coli* in participants without known risk factors from the southeastern United States

**DOI:** 10.1101/2024.02.03.24302254

**Authors:** Coralis Rodriguez Garcia, William A. Norfolk, Amanda K. Howard, Amanda L. Glatter, Megan S. Beaudry, Nicholas A. Mallis, Michael Welton, Travis C. Glenn, Erin K. Lipp, Elizabeth A. Ottesen

**Author notes:** Center for Disease Control and Prevention, Atlanta, Georgia, USA. Department of Epidemiology of Microbial Diseases, Yale School of Public Health, New Haven, CT, USA.

## Abstract

We evaluated gut carriage of extended spectrum beta lactamase producing *Enterobacteriaceae* (ESBL-E) in southeastern U.S. residents without recent in-patient healthcare exposure. Study enrollment was January 2021-February 2022 in Athens, Georgia, U.S. and included a diverse population of 505 adults plus 50 child participants (age 0-5). Based on culture-based screening of stool samples, 4.5% of 555 participants carried ESBL-Es. This is slightly higher than reported in studies conducted 2012-2015, which found carriage rates of 2.5-3.9% in healthy U.S. residents.

All ESBL-E confirmed isolates (n=25) were identified as *Escherichia coli*. Isolates belonged to 11 sequence types, with 48% classified as ST131. Ninety six percent of ESBL-E isolates carried a *bla_CTX-M_* gene. Isolated ESBL-Es frequently carried virulence genes as well as multiple classes of antibiotic resistance genes. Long-term colonization was common, with 64% of ESBL-E positive participants testing positive when rescreened three months later. One participant yielded isolates belonging to two different *E. coli* sequence types that carried *bla_CTX-M-1_* genes on near-identical plasmids, suggesting intra-gut plasmid transfer.

Isolation of *E. coli* on media without antibiotics revealed that ESBL-*E. coli* typically made up a minor fraction of the overall gut *E. coli* population, although in some cases they were the dominant strain. ESBL-E carriage was not associated with a significantly different stool microbiome composition. However, some microbial taxa were differentially abundant in ESBL-E carriers. Together, these results suggest that a small subpopulation of US residents are long-term, asymptomatic carriers of ESBL-Es, and may serve as an important reservoir for community spread of these ESBL genes.

**Importance:** Antibiotic resistant bacteria, especially *Enterobacteriaceae* carrying ESBLs, have become an increasing public health threat. Increasing numbers of community-associated infections (47% of ESBL-E infections in the U.S.) in participants without healthcare exposure is particularly concerning. This study found that 4.5% of a southeastern United States study population, without in-patient healthcare exposure, were asymptomatically colonized with ESBL-E, and 64% of ESBL-E positive participants were still positive when rescreened 3 months later. This suggests that the gut microbiome of healthy participants may represent an understudied community reservoir of ESBL genes and ESBL *Escherichia coli* in the U.S.

## Introduction

Antimicrobial resistance is a serious and growing public health threat worldwide (1, 2) that contributes to increased complication rates, as well as increased treatment costs (3, 4). Extended-spectrum beta-lactamase (ESBL) producing *Enterobacteriaceae* (ESBL-E) are listed as a serious threat by the U.S. Center for Disease Control and Prevention (CDC), contributing to 197,400 infections and 9,100 deaths in 2019 (2).

ESBL-E were first reported in 1983 and have since spread rapidly throughout the world (5–8). ESBL enzymes confer resistance to multiple antibiotics including penicillin, monobactams, and cephalosporins, commonly used to treat infections caused by Gram negative bacteria (9). There are multiple classes of ESBL enzymes including TEM, SHV, OXA and CTX-M (10–12). Recently, attention has been drawn to the CTX-M class ESBLs as it is currently the most common ESBL found worldwide (13–16). CTX-M-15 is typically associated with *E. coli* sequence type 131 (ST131), a frequent causative agent of extraintestinal infections and outbreaks (17, 18). The successful dissemination of this *E. coli* clone contributed to the wide spread of this ESBL enzyme (19). Another factor contributing to the rapid dissemination of ESBLs is their frequent association with mobile elements including plasmids (20–22). Microorganisms often carry ESBL genes in plasmids from the groups IncF, IncI1, IncA/C and IncHI2, facilitating horizontal gene transfer of antibiotic resistance (23). These plasmids also carry antibiotic resistance genes for other classes, easily resulting in multi-drug resistant organisms (24).

In a meta-analysis by Bezabih et al. (25), the global intestinal carriage rate of ESBL-*E. coli* in the community increased 10-fold during 2001-2020, highlighting the significance of investigating community-associated ESBL-E. The global prevalence of ESBL-E fecal carriage in the community is highly variable throughout the globe, with substantially higher rates (76.3%) in Tanzania and the lowest (1.9%) in Australia (25, 26). The exact causes of these geographic variations are unknown, although they have been associated to factors including the use of antibiotics in food animals and sanitation standards (27).

Few studies have examined ESBL-E carriage in healthy individuals in the U.S. Doi et al. (28, 29) reported a 3.9% prevalence of ESBL-*E. coli* in community-associated samples from outpatient clinics in 5 U.S. states collected in 2009-2010. Vento et al. (30) found that only 1 of 101 healthy U.S. military personnel based in the U.S. (May-June 2011) carried an ESBL-E. Weisenberg et al. (31), reported a 2.5% colonization rate for ESBL-E in 2012, among New York residents in participants at pre-travel or no international travel planned. Finally, Islam et al. (32) reported an ESBL-E carriage rate of 3.5% among stool samples from healthy children collected 2013-2015 in three U.S. cities. Overall, these studies suggest that carriage of ESBL-E in the U.S. is low. However, the steady increase of community-associated ESBL-E infections from 2013-2019 (2, 29) suggest a need for updated data on ESBL-E community carriage in the U.S.

The aim of this work, was an examination of the prevalence and risk factors for gut colonization of ESBL-E in the community among participants living in or near Athens, Georgia (GA), U.S. We also evaluated the frequency of long-term (∼3 month) carriage of ESBL-E among positive participants. Finally, we performed an in-depth genomic analysis, including mobile elements, of ESBL-E isolated in this study and a comparison of gut microbiome community composition between carriers and non-carriers.

## Results

We recruited a total of 555 participants including 505 adults and 50 children between January 2021 and November 2021 from the southeastern U.S. (mostly northeastern Georgia) (Fig. S1). Overall, the study population reflected the demographic composition of the study area as expected based on U.S. census data (Table S2), with some over-representation of participants who identified as white and non-Hispanic, were 18-39 years of age, and/or reported female sex. (Table S1).

### Carriage of ESBL *Enterobacteriaceae* (ESBL-E)

From 555 samples, 25 participants (4.5%) were positive for stool carriage of an ESBL-E. One of the 25 ESBL-E positive participants was a child, whose parent also tested positive. Of the 25 ESBL-E positive participants, 22 provided a second sample at follow-up (97-176 days later), including the parent-child dyad. 14 of those 22 samples (64%) were ESBL-E positive, whereas the remaining 8 were ESBL-E negative.

ESBL-E carriage was not identified as significantly associated with any of the demographic or socioeconomic risk factors examined (Table 1, Table S1). We observed slightly, but not significantly, increased incidence of ESBL-E in participants identified as biological males and Asian or Black/African American race identities. The full list of occupational, lifestyle, and environmental risk factors tested is available in supplemental table S1. A subset of the study population regularly experienced interaction with increased risk environments; however, these exposures were not significantly associated with ESBL-E carriage. ESBL-E carriage was also not associated with significant differences in self-reported gastrointestinal (GI) distress, urinary tract infection, or antibiotic usage. A review of the health information provided by ESBL-E positive participants did not suggest severe chronic health problems.

**Table 1:**
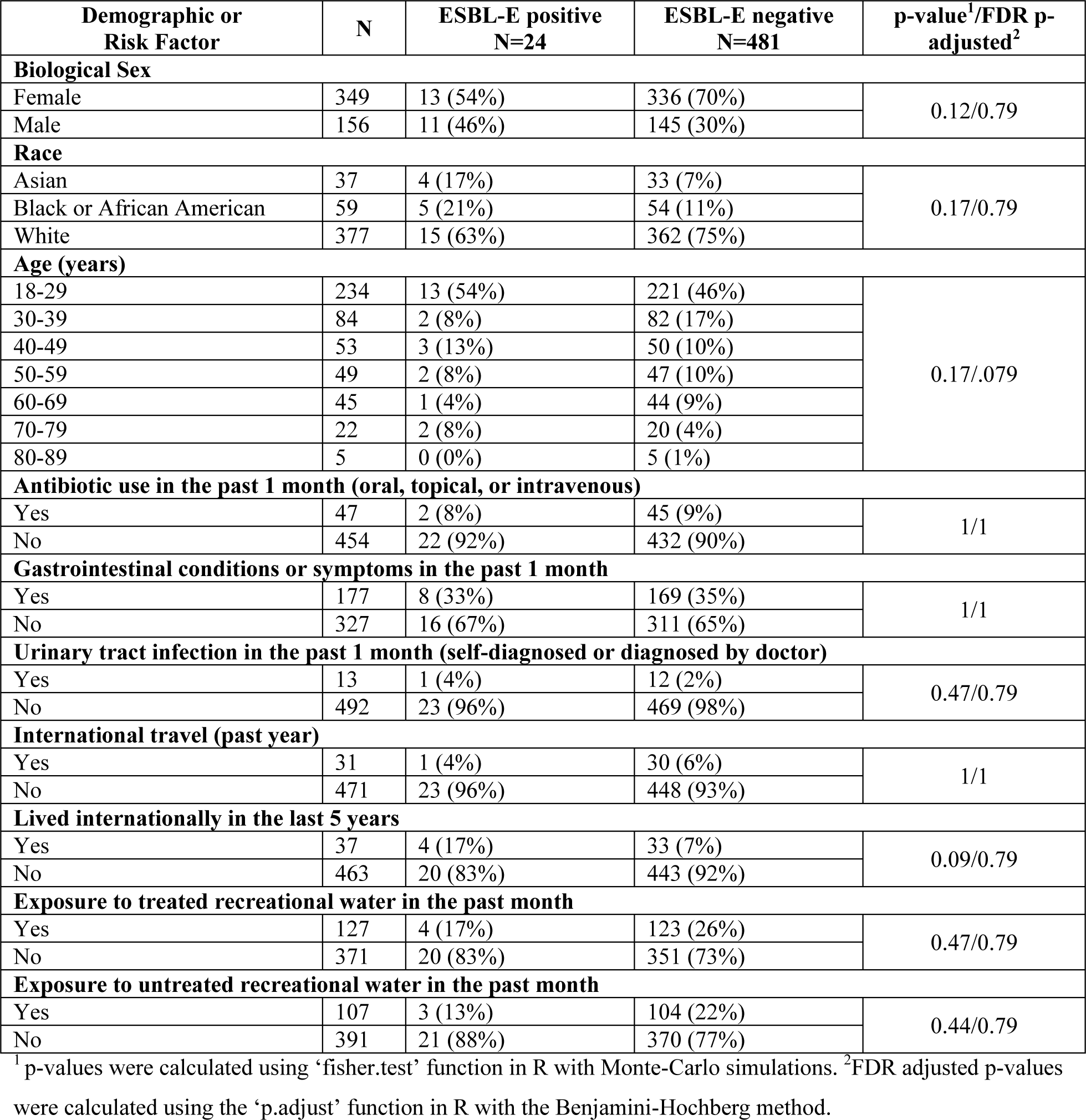
Selected demographic characteristics, risk factors, and presence of ESBL-E among all adult participants. The demographic data presented on Table 1 are based on the questionnaire responses from the first visit of all adult participants. The child demographics were not included in the tables or statistical analyses of risk factors, as they were not considered independent of the parent participants.

| Demographic or Risk Factor | N | ESBL-E positive<br>N=24 | ESBL-E negative<br>N=481 | p-value<sup>1</sup>/FDR p-adjusted<sup>2</sup> |
| --- | --- | --- | --- | --- |
| **Biological Sex** |  |  |  |  |
| Female | 349 | 13 (54%) | 336 (70%) | 0.12/0.79 |
| Male | 156 | 11 (46%) | 145 (30%) |  |
| **Race** |  |  |  |  |
| Asian | 37 | 4 (17%) | 33 (7%) | 0.17/0.79 |
| Black or African American | 59 | 5 (21%) | 54 (11%) |  |
| White | 377 | 15 (63%) | 362 (75%) |  |
| **Age (years)** |  |  |  |  |
| 18-29 | 234 | 13 (54%) | 221 (46%) | 0.17/.079 |
| 30-39 | 84 | 2 (8%) | 82 (17%) |  |
| 40-49 | 53 | 3 (13%) | 50 (10%) |  |
| 50-59 | 49 | 2 (8%) | 47 (10%) |  |
| 60-69 | 45 | 1 (4%) | 44 (9%) |  |
| 70-79 | 22 | 2 (8%) | 20 (4%) |  |
| 80-89 | 5 | 0 (0%) | 5 (1%) |  |
| **Antibiotic use in the past 1 month (oral, topical, or intravenous)** |  |  |  |  |
| Yes | 47 | 2 (8%) | 45 (9%) | 1/1 |
| No | 454 | 22 (92%) | 432 (90%) |  |
| **Gastrointestinal conditions or symptoms in the past 1 month** |  |  |  |  |
| Yes | 177 | 8 (33%) | 169 (35%) | 1/1 |
| No | 327 | 16 (67%) | 311 (65%) |  |
| **Urinary tract infection in the past 1 month (self-diagnosed or diagnosed by doctor)** |  |  |  |  |
| Yes | 13 | 1 (4%) | 12 (2%) | 0.47/0.79 |
| No | 492 | 23 (96%) | 469 (98%) |  |
| **International travel (past year)** |  |  |  |  |
| Yes | 31 | 1 (4%) | 30 (6%) | 1/1 |
| No | 471 | 23 (96%) | 448 (93%) |  |
| **Lived internationally in the last 5 years** |  |  |  |  |
| Yes | 37 | 4 (17%) | 33 (7%) | 0.09/0.79 |
| No | 463 | 20 (83%) | 443 (92%) |  |
| **Exposure to treated recreational water in the past month** |  |  |  |  |
| Yes | 127 | 4 (17%) | 123 (26%) | 0.47/0.79 |
| No | 371 | 20 (83%) | 351 (73%) |  |
| **Exposure to untreated recreational water in the past month** |  |  |  |  |
| Yes | 107 | 3 (13%) | 104 (22%) | 0.44/0.79 |
| No | 391 | 21 (88%) | 370 (77%) |  |

<sup>1</sup>p-values were calculated using 'fisher.test' function in R with Monte-Carlo simulations. <sup>2</sup>FDR adjusted p-values were calculated using the 'p.adjust' function in R with the Benjamini-Hochberg method.

International travel has been widely associated with ESBL colonization (33). The proportion of ESBL-positive participants that lived internationally in the last 5 years is higher than the ESBL-negative participants (17% vs. 7%); however, it was not significantly associated with ESBL-carriage (fdr-corrected p-value 0.09). Only 8 participants (all ESBL-negative) reported international travel in the previous 30 days prior to sample collection.

### Characteristics of ESBL-E isolates

#### Genome assemblies and phylogenetic relationships

All confirmed ESBL-E were identified as *Escherichia coli* by MALDI-TOF. High-quality draft genome sequences were obtained for the ESBL-E isolates of each 25 initial and 14 second visit samples (Table S3). Genome sizes ranged from 4,981,979 bp to 5,465,567 bp.

Isolates belonged to phylogroups B2 (17 isolates), D (4), A (2) B1 (2) and F (1). Of the B2 isolates, 12 belonged to the uropathogenic group ST131, including members with *fimH*41 and *fimH*30 alleles (Fig. S2). Other closely related groups, ST2279 (2 isolates), ST1193 (2) and ST636 (1) were also present (Fig. 1F). All the samples from second visits that yielded confirmed ESBL-E positive were near-clonal (3-55 SNPs) with the isolate of the initial visit from that participant with the sole exception of the isolates from participant 497 (Fig. 1E). The initial isolate from participant 497 was assigned to ST154 in phylogroup B1, whereas the isolate from this participant’s second sample, 497R, was assigned to ST106 in phylogroup D (Fig. 1A-F). The parent-child dyad samples were both assigned to phylogroup A but had different sequence types: ST10 (parent) and ST305 (child) and 22,309 SNP differences.

**Figure 1:**
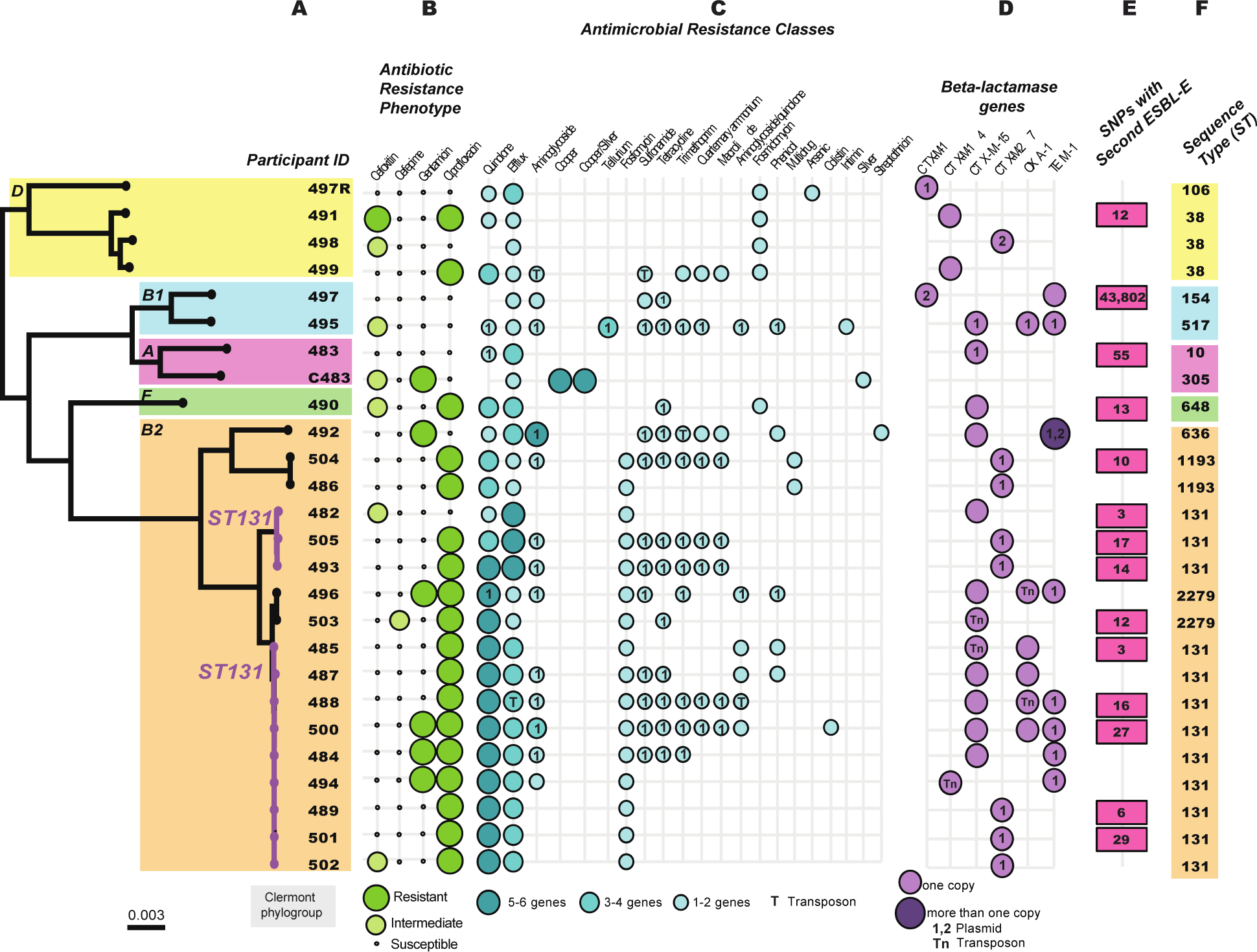
Characteristics and classification of ESBL-E isolates. A) Core-genome single nucleotide polymorphisms (SNPs) phylogenetic tree of ESBL-E isolates per participant colored by Clermont phylogroup, highlighting ST131 isolates. B) Phenotypic characterization of antibiotic resistance by isolate tested in Sensititre, after the confirmation as ESBL-E given their resistance to beta-lactams: Cefotaxime and Ceftazidime. C) Antimicrobial resistance genes found by AMRFinderPlus in the genome, transposons (T) or plasmids (1) of each isolate classified by antimicrobial class. D) Beta-lactamase genes found in each isolate, chromosomally or in mobile elements. Circles labelled with a number match plasmids described in Table S5 for each isolate. E) SNPs between first and second sample confirmed as ESBL-positive to determine clonality of isolates. F) Sequence type (ST) classification of each isolate, colored by phylogroup. Second samples collected (R) are not included in the figure as they were the same sequence type (ST) and share almost the same genetic content as their original sample, with the exception of 497R.

#### Antibiotic resistance profile

All confirmed ESBL-E positive isolates were phenotypically resistant to Ampicillin, Cefotaxime, Ceftazidime, Cefpodoxime, Ceftriaxone, Cefazolin, and Cephalothin. They presented variable resistance to Cefoxitin and Cefepime as shown in Figure 1B. Of the 25 initial ESBL-E isolates, 18 had resistance to Ciprofloxacin (72%) and 6 were resistant to Gentamicin (24%). ESBL-E isolates from the second visit exhibited the same phenotypic antibiotic resistance profile as their original sample except for 491R, which showed a decreased resistance to Cefoxitin compared to the isolate 491. Some second-visit isolates (C483R and 487R) had resistance to Cefoxitin but were susceptible to other beta-lactams tested and were not confirmed as ESBL-E. None of the isolates were resistant to the carbapenems tested.

All isolates, except for the child sample, carried at least one *bla_CTX-M_*beta lactamase gene (Fig. 1D). The most predominant beta lactamase gene was *bla_CTX-M-15_* in 12 isolates, followed by *bla_CTX-M-27_* in 8 isolates, *bla_CTX-M-14_* in 3 isolates, and *bla_CTX-M-1_*in 2 isolates. *bla_CTX-M-15_* was located on a plasmid in 2 isolates and located within a chromosomally encoded transposon in 2 isolates. *bla_CTX-M-15_*was chromosomally encoded without an obvious mobile genetic element in the remaining 8 isolates (Fig. 1D). All isolates with *bla_CTX-M-27_* carried the gene in IncF plasmids (Table 2).

**Table 2:**
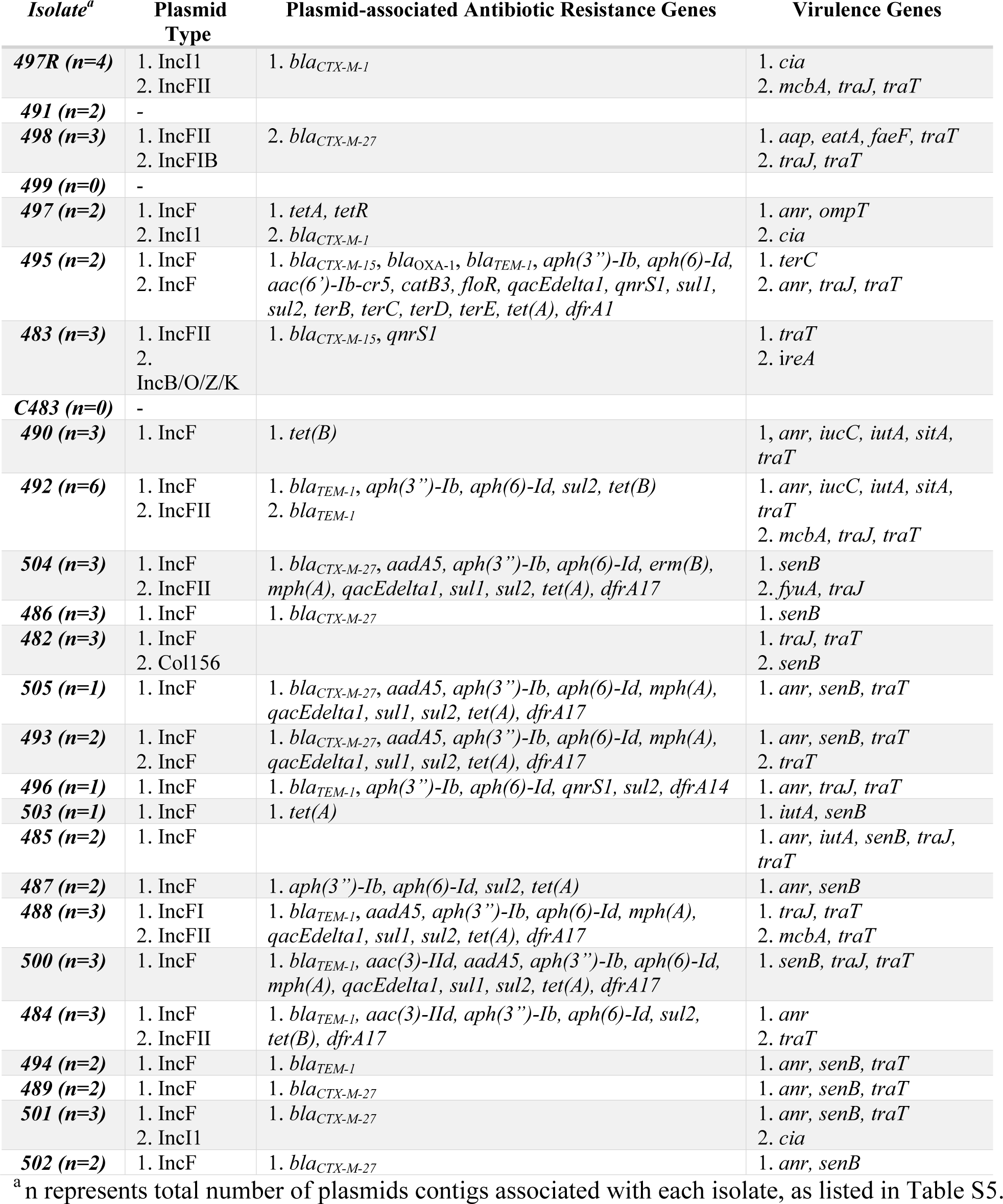
Assembled plasmids carried by each isolate identified by PlasmidFinder with encoded antibiotic genes identified by AMRFinderPlus and encoded virulence genes identified by VirulenceFinder. Plasmid number in each isolate matches the numbers in panels C and D of Figure 1 and Figure 2.

Other beta lactamase genes identified in ESBL-E isolates included *blaEC*, *bla_TEM-1_*, and *bla_OXA-1_*. All ESBL isolates carried the *blaEC* gene that confers resistance to Ampicillin, except for isolates 504 and 486, which carried *blaEC5* (not shown). Beta lactamase genes found in lower prevalence were *bla_TEM-1_* (carried by 8 isolates) and *bla_OXA-1_* (carried by 6 isolates). Seven copies of *bla_TEM-1_* were carried by IncF plasmids, and one isolate had a copy in the chromosome. *bla_OXA-1_* was encoded in an IncF plasmid of one isolate, in transposons of two isolates and chromosomally in three isolates (Fig. 1D). Isolate 492 carried three copies of *bla_TEM-1_*, two of them in two different plasmids and one chromosomally. No currently characterized ESBL beta lactamase genes were identified in the child sample, C483. It is not currently clear whether this isolate carries an uncharacterized gene or may have lost its ESBL gene following initial isolation on selective media.

In addition to these beta lactamases, all isolates carried antimicrobial resistance genes in the efflux class, while 16 carried AR genes for Fosfomycin, 12 for sulfonamides, 12 for tetracycline, 10 for trimethoprim, 7 for macrolides, 8 for quaternary ammonium and 5 for phenicol (Fig. 1C). Interestingly, the child isolate C483 carried two operons (*pco* and *sil)* for resistance genes in metal classes like Copper and Silver, both in close association with transposase genes. Nearly all isolates (22 of 26, including 497R) carried quinolone resistance genes. All ST131 isolates had mutations on *gyrA* and *parE* genes and exhibit Ciprofloxacin resistant phenotype, except isolate 482 (Fig. S2). For participants with a confirmed ESBL-E isolate from their second visit, all but two encoded the same resistance genes on re-isolation. One of these is 497R, which as discussed elsewhere, denoted a different sequence type from the initial visit isolate. The other, 493R, was identified as clonal (14 SNPs) but lacked erythromycin, trimethoprim, streptomycin, tetracycline, and sulfamethoxazole antibiotic resistance genes that were present in the original isolate, 493. Both isolates carried a near-identical IncF plasmid, but in 493 this plasmid contained an IS6 insertion element that encoded the resistance genes and is missing in the plasmid carried by 493R (possibly lost in a deletion event) (Fig. S3).

#### Virulence genes and Plasmids

Phylogroup B2 isolates carried a large variety of virulence genes associated with extraintestinal *E. coli* (ExPEC) and had three or more genes associated with sepsis-associated *E. coli* (SEPEC) (Fig. 2).

**Figure 2:**
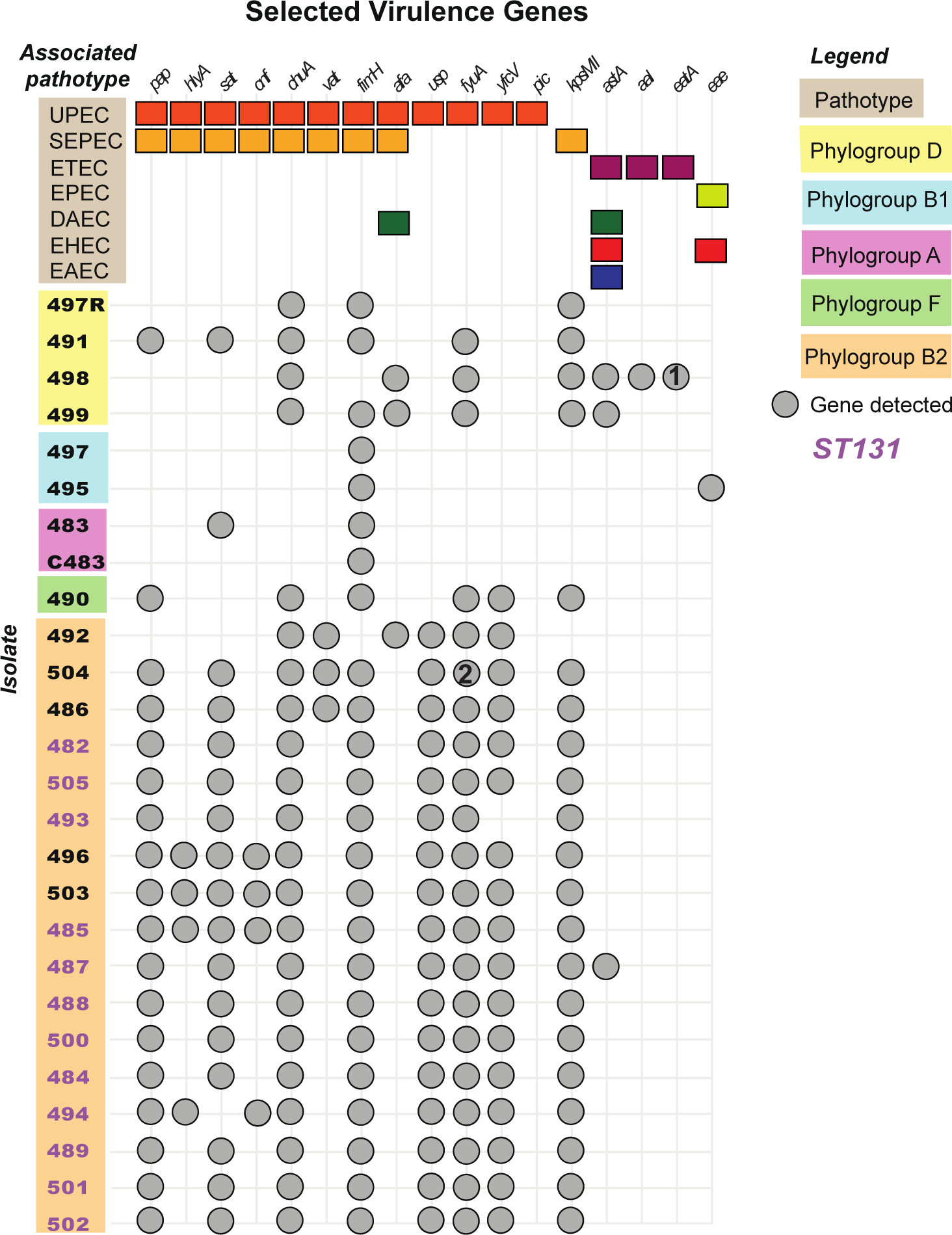
Selected virulence genes associated with pathogenic *E. coli* pathotypes. Virulence genes were identified by VirulenceFinder using the genome assemblies of each isolate. Only selected virulence genes are shown based on their associated pathotype. Colored boxes represent the genes associated with each pathotype and gray circles indicate the presence of that gene in each isolate. Isolates are clustered by Clermont phylogroups matching Figure 1. Circles labelled with a number match plasmids described in Table S5 for each isolate.

However, virulence genes were not limited to phylogroup B2 isolates. The *afa* genes, encoding for afimbrial adhesins, are also associated with diffusely adherent *E. coli* (DAEC) and were found in isolates 498 and 499 from phylogroup D and 492 from phylogroup B2. Six of 9 genes in the adhesin associated locus (*aal*) that encodes for the Coli surface antigen 23 (CS23) were present in isolate 498, which also carried the *eatA* gene, encoding for ETEC autotransporter A. Isolate 495 carried the epsilon subtype of intimin outer membrane protein gene, *eae*, associated with enteropathogenic *E. coli* (EPEC), hemorrhagic *E. coli* (EHEC) and Shiga-toxin *E. coli* (STEC). Isolate 488 has 3 P fimbriae genes (A, C, F) in the same transposon as *bla_OXA-1_*in addition to other resistance genes (*aac(6’)-Ib-cr5*, *ermD*, *mdtM*).

The most frequently identified and biologically significant plasmids identified belonged to the IncF group, with 24 of the 26 unique ESBL-E isolates carrying at least one plasmid in this group. Most beta lactamases genes located within plasmids were carried by IncF plasmids, except for isolates 497 and 497R which carried *bla_CTX-M-1_*in an IncI1 plasmid (Table 2). The only plasmid carrying more than one beta-lactamase gene was the IncF in isolate 495, which carried genes *bla_TEM-1_*, *bla_CTX-M-15_*, and *bla_OXA-1_*. Isolate 492 has 2 plasmids each carrying a copy of *bla_TEM-1_*.

Eight isolates carried IncF plasmids that encoded *bla_CTX-M-27_*, all of which also carried the plasmid-encoded enterotoxin, *senB*. *bla_CTX-M-27_* was encoded in an IS6 transposon array frequently associated with up to 16 additional antibiotic resistance genes (Fig. S4). Two of these isolates, 486 and 504, were near-clonal (52 SNPs) and carried plasmids that were 99.98% identical, with the main difference being two 65Kb IS6 transposon arrays encoding (among other genes) 16 resistance genes that are present in the plasmid carried by isolate 504 but absent in 486 (Fig. S5).

Three isolates carried an IncI1 plasmid, including 497 and 497R. Isolates 497 and 497R, from the same participant at different times and in different sequence groups, shared a near-identical (99.7% pairwise identity) plasmid encoding *bla_CTX-M-1_*(Fig. S6). Plasmid group IncY was identified in 2 isolates, while replicons for IncI2, IncH, IncB/O/K/Z and IncN were each present in one isolate accompanied by an IncF plasmid (Table 2).

### Carriage of antibiotic resistant *E. coli* among ESBL-E positive participants

We also examined the overall prevalence of AR among commensal *E. coli* in ESBL-E positive participants. Up to 48 *E. coli* isolates were tested for resistance to ampicillin, ceftriaxone, ciprofloxacin, tetracycline, and trimethoprim. Participants 500 and 498 yielded no *E. coli* colonies on this medium. Six of 25 ESBL-E positive participants showed dominance of beta-lactam resistant strains in their commensal *E. coli* community, with more than 50% of the colonies resistant to both ceftriaxone and ampicillin (Fig. 3). Five participants carried commensal *E. coli* resistant to ciprofloxacin. High levels (>50% colonies) of tetracycline resistance were found in 10 of the 25 ESBL-E positive participants. Trimethoprim resistance was less common, with only six participants showing high resistance (>50% colonies) (Fig. 3). In the second samples provided, eight participants had an increase in the proportion of *E. coli* colonies resistant to both ceftriaxone and ampicillin. Most of the second samples that were ESBL-E negative, had a decreased proportion of commensal *E. coli* colonies that were resistant to the antibiotics tested.

**Figure 3:**
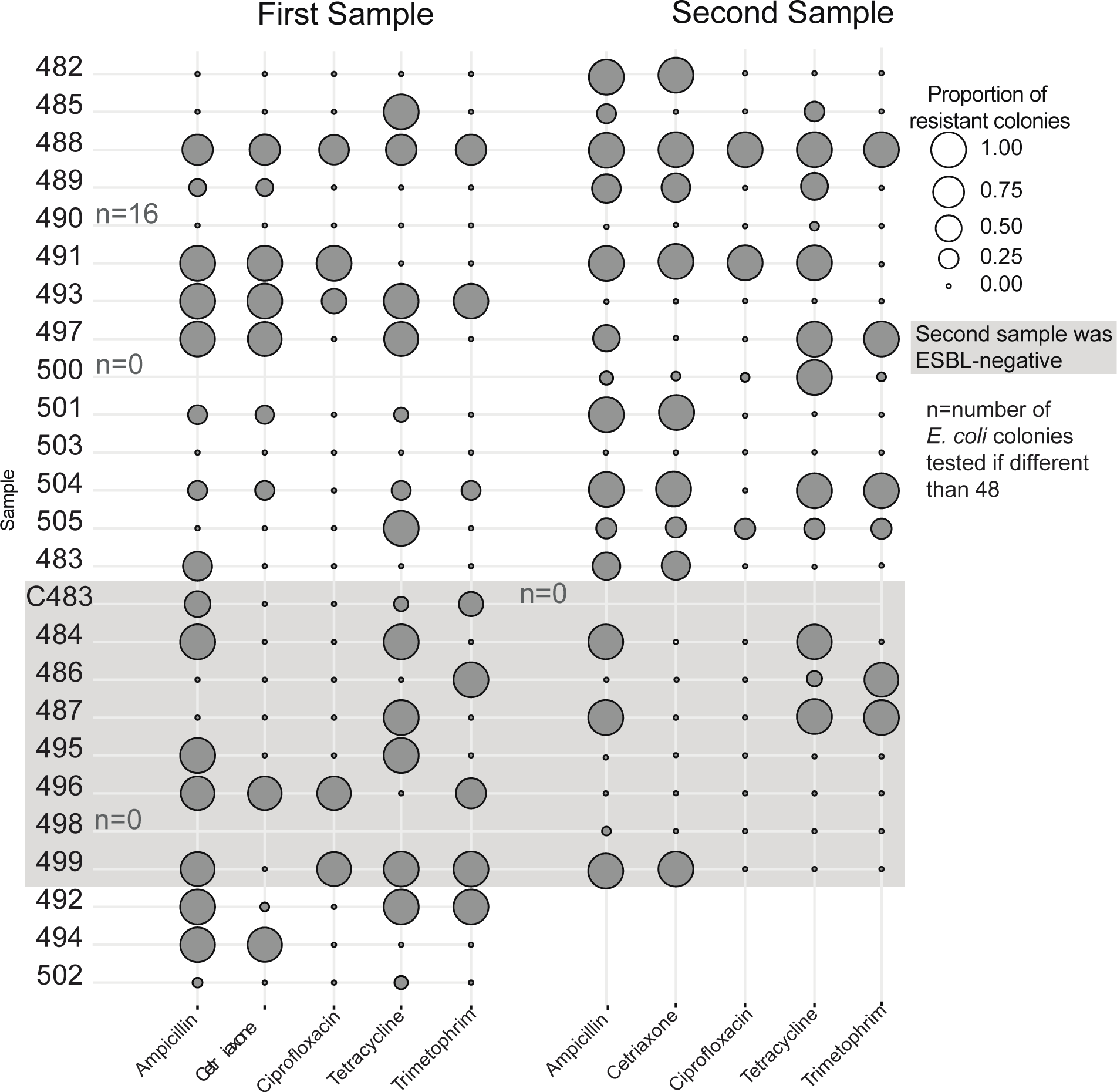
Antibiotic resistance phenotype of commensal *E. coli* isolated from ESBL-E positive participants. A range between 1-48 *E. coli* colonies isolated from ESBL-E positive participants were patched into Mueller Hinton II agar with antibiotics at CSLI standard levels. Each circle size and color intensity show the proportion of those colonies that were resistant to the antibiotic tested.

#### Gut microbial community composition in ESBL-E positive and ESBL-E negative participants

ESBL-E colonization was not associated with significantly different stool microbiome alpha diversity (Fig. S7). Stool microbiome composition as evaluated by weighted Bray-Curtis distances (Fig. 4A) resulted in a significant PERMANOVA p-value of 0.03. However, the R^2^ value was 0.003, indicating that ESBL-E carriage explained only a very small proportion of community variance. Participants with negative second visit samples did not exhibit significantly larger between-sample shifts in stool microbiome composition than participants with continued colonization at re-sampling (Fig. 4B; Fig. S8).

**Figure 4:**
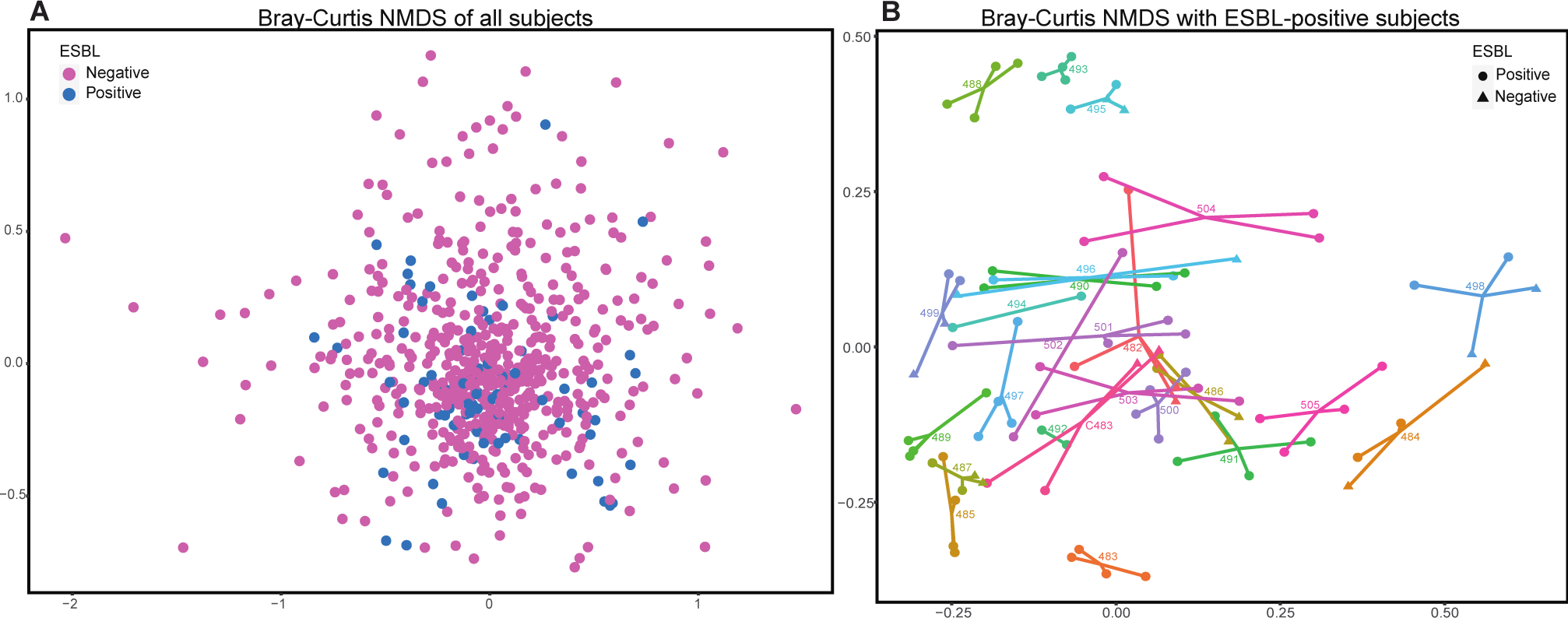
Microbial community similarities based on ESBL-E carriage. A) Bray-Curtis distances of metagenomic microbiome samples from all participants, organized in a non-metric multidimensional scaling (NMDS). Magenta dots represent participants that were ESBL-E negative while blue dots represent participants colonized with ESBL-E. B) Comparison of samples (first, duplicate of first, second and duplicate of the second sample) from the same ESBL-E positive participant, calculated by Bray-Curtis distances. Each number indicates the participant ID, while circles or triangles represent the ESBL-E status by sample.

The relative abundance of most microbial classes was similar between ESBL-E positive and ESBL-E negative samples (Fig. S9). However, DESeq2 analysis identified 21 amplicon sequence variants (ASVs) that were significantly enriched and 50 that were significantly depleted in samples from ESBL-E positive vs. ESBL-E negative participants (Fig. 5).

**Figure 5:**
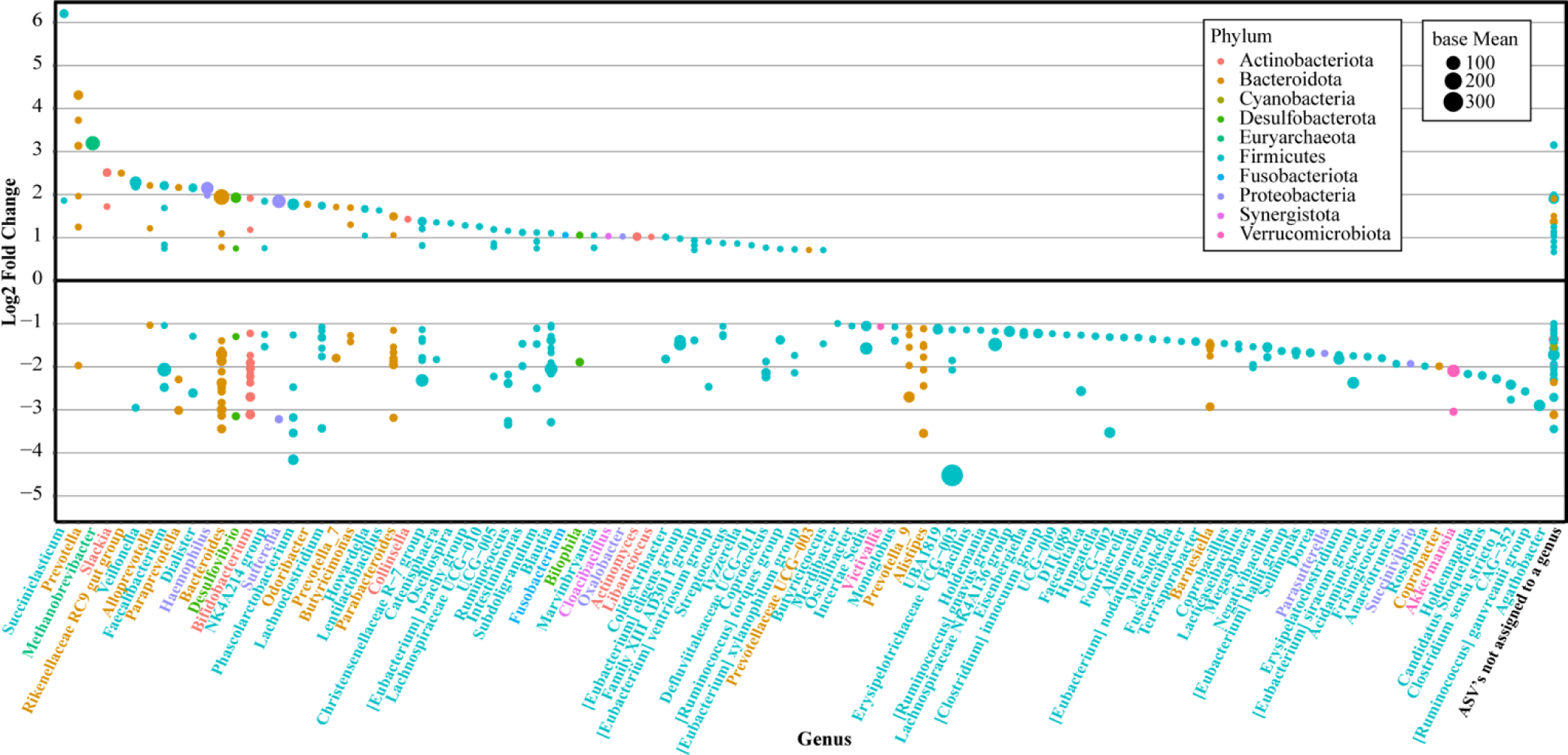
Genus assignments for ASVs identified as enriched or depleted in ESBL-E positive participants. ASVs identified as significantly enriched or depleted by DESeq2 analysis with local fit and Wald test identifying significantly different ASVs under an alpha=0.001. ASVs are colored by phylum and organized by genus. Point size shows normalized mean counts and y axis shows log2fold change in ESBL-E positive participants. ASVs with a positive log2fold change were enriched, while ASVs with a negative log2fold change were depleted in ESBL-E positive participants.

## Discussion

A key goal of this work was to evaluate asymptomatic carriage of ESBL-E in U.S. residents without significant healthcare exposure. We observed a carriage rate of 4.5% in our study population, which was recruited from the vicinity of the city of Athens, in northeastern Georgia. This reflects a higher prevalence than previous reports from the U.S. that range between 1.7% and 3.5% (26, 30, 32), although this remains lower than carriage rates reported from European and African countries (34–36). One caveat is that our study participants were recruited from the southeastern U.S., which has reported a higher rate (19.9%) of healthcare associated ESBL-E infections than the national average of 12.2% (37).

Our study did not identify any demographic, socioeconomic, environmental, or health-associated risk factors that were significantly associated with the carriage of ESBL-E. This contrasts with previous studies that have reported antibiotic usage (38) and recent international travel (39) as correlated to ESBL-E carriage. These results may have been impacted by the COVID-19 pandemic, which limited international travel during the study period (January 2021-February 2022). This lack of association with specific risk factors suggests that ESBL *E. coli* may be circulating, albeit at low rates, among the general population in the study area.

A large fraction (14 of 22) of ESBL-E positive participants remained positive when re-tested at least 3 months after their initial visit, which is similar to previous reports in Sweden (40). That study, as well as others, found that sequence type ST131 or strains classified in the B2 and D phylogroups were more likely to persist (40, 41). However, our persistent isolates were distributed across many *E. coli* phylotypes, suggesting that persistent colonization is not confined to these phylogroups (42).

Overall, the most prevalent group of ESBL *E. coli* was B2, which matches previous studies in North America (43–46). In terms of sequence types, 46% of the isolates were identified as belonging to ST131, a globally distributed uropathogenic clade that is widely associated with ESBL gene carriage and identified as the most frequent multidrug resistant extraintestinal pathogenic *E. coli* (47). ST131 subclades C1, C2 and C1-M27 (as classified by *bla_CTX-M_* and *fimH* (48)) were all present.

All but one isolate carried at least one gene encoding for an CTX-M type enzyme, for a total prevalence of 96%, compared to the 90% reported in a previous review of community isolates from different geographical regions worldwide (49). This suggests that *bla_CTX-M_*carrying *E. coli* may be a key driver of community ESBL-E spread in the southeastern U.S. In fact, an *E. coli* carrying *bla_CTX-M-15_*has been isolated from a stream sample in Athens, GA (50). Among the isolates that carried *bla*_CTX-M_ genes, *bla_CTX-M-15_* was the most predominant, followed by *bla_CTX-M-27_* and *bla_CTX-M-14_*. This agrees with previous reports that *bla*_CTX-M-15_ is the most abundant beta lactamase gene circulating in the U.S., closely followed by *bla*_CTX-M-27_ (51, 52), although a third study of urinary tract infections from gram-negative pathogens isolated in hospitals of Canada and the U.S. (2010-2014) reported *bla_CTX-M-14_* as more abundant than *bla_CTX-M-27_*(53). *bla_CTX-M-27_* was primarily associated with ST131 C1-M27 and C1/H30R clades as previously reported (54) but was also present in ST131 clones with *fimH41*. In all recovered isolates with this gene, the *bla_CTX-M-27_* was borne on an IncF-type plasmid.

In addition to the resistance shown to beta-lactams antibiotics, 72% of the ESBL-E isolates showed resistance to Ciprofloxacin and 24% were resistant to Gentamicin. Ciprofloxacin resistance is widely associated with ST131 (17); however, 7 isolates from other groups also exhibited ciprofloxacin resistance suggesting a broader relationship. A previous study of extra-intestinal pathogenic ESBLs from hospitalized patients in India reported a slightly lower prevalence of 65% (55). It remains unclear whether this increased prevalence is because ciprofloxacin resistance is more common in gut isolates or whether this is due to the population studied.

The most abundant plasmid type found in our isolates was IncF, which also was the plasmid type carrying the most antibiotic resistance and virulence genes, as has been widely reported (56). While plasmids from the incompatibility group IncI1 are typically associated with transfer of beta lactamase genes (57), we only isolated ESBL-carrying IncI1 plasmids from two participants. The IncI1 plasmid from participant 497, is of particular interest because the ESBL-*E. coli* isolates obtained from the first and second fecal samples provided by this participant carried a near-identical copies of this plasmid but belonged to two different lineages of *E. coli*. This suggests a recent transfer of this plasmid in the human gut. Similar events have been reported elsewhere (58–61) and have been used to argue for the potential role of the human gut as a key site of horizontal gene transfer of antibiotic resistance genes.

In agreement with previous studies (62), many of our isolates, particularly those belonging to the B2 phylogroup, carried virulence genes associated with extra-intestinal pathogenic *E. coli* including uropathogenic (UPEC) and sepsis associated (SEPEC) strains. The *fimH* gene was the most prevalent virulence factor, followed by UPEC-associated genes *chuA, fyuA, usp*, and *yfcV* and *kpsMII* (found in 84% of ESBL-E isolates). Other virulence genes frequently found in our study include *iutA*, *papA*, *papC*, *sat*, *vat*, *cnf1* and *hlyA*, as previously shown by Iseppi et. al (63), however in contrast to their results *sfa* was not found in any isolate and *afa* genes were only in 3 isolates. Remarkably, the *eae* gene, previously used to classify EPEC isolates (64), was found in the isolate from participant 495, who also carried the only plasmid in this study with 3 different beta-lactamase genes. The presence of virulence genes in plasmids could explain the hybrid classification into different pathotypes of *E. coli* and contribute to the evolution of these pathotypes by accumulating virulence factors within the commensal community (65–67).

We also examined the frequency of antibiotic resistance among the overall population of commensal *E. coli* present in stool samples from ESBL-E positive participants. More than half of ESBL-E positive participants carried ampicillin- (64%) and tetracycline- (56%) resistant *E. coli*. However, presumptive ESBL-E made up most commensal *E. coli* in initial samples from only 6 of 25 ESBL-positive participants. This agrees with previous studies that in most cases the ESBL-producing *E. coli* is not the dominant strain among gut *E. coli* (36). Interestingly, 8 of the 22 second samples showed an increase in the fraction of presumptive ESBL-*E. coli* isolated which could indicate a highly dynamic population structure and/or possible genetic exchange of antibiotic resistance genes among commensal *E. coli*.

Alpha and beta diversity analysis suggests that ESBL-E carriage was not associated with substantially different gut microbiome composition or diversity, consistent with the results of other studies (68, 69). However, pairwise analysis identified multiple taxa with significantly different abundance in carriers and non-carriers. Similar to other studies, we observed enriched taxa belonging to *Prevotella* (69, 70) and depletion of taxa belonging to *Sellimonas* (71) and *Bacteroides uniformis* (72). On the other hand, some taxa that other groups found enriched in ESBL-E carriers were identified as depleted in our ESBL-E communities, including *Clostridiales* (69), *Erysipelotrichaceae*, *Lactococcus*, *Bilophila* and *Negativibacillus* (70). In addition, *Desulfovibrio* and *Oscillospira* genera were identified as enriched among ESBL-E carriers in our study but depleted in another (68). These differences could be explained by the species of ESBL-*Enterobacteriaceae* in each study, given that the microbial population differs based on the ESBL-E species (71), or other factors influencing the microbiome composition as discussed before (73).

Notable limitations of our study include that it was restricted to a specific geographic range (Athens, GA and vicinity) and that the study commenced during the global COVID-19 pandemic, which caused multiple large-scale changes in behavior including limitations on international travel. In addition, there was a possible bias provided by the participant self-reporting of risk factors. However, overall our results suggest that a subset of southeast U.S. residents are likely asymptomatic carriers of ESBL-*E. coli*.

## Conclusions

To our knowledge this is the first genomic analysis of community associated ESBL-E carriage in the southeastern U.S. including long-term colonization, as previous studies only focused on the prevalence of ESBL-E or on healthcare-associated isolates. If the results from this study can be extrapolated, it suggests the potential for a small but notable increase in ESBL-E carriage in the U.S. since 2015, consistent with reports that the frequency of community-acquired ESBL-E infections also increased over this period (25) and supporting the role community-associated isolates in the incidence of ESBL-E outbreaks. Long term (>3 months) colonization was common in the study population, which underscores the potential of the human gut microbiome to serve as a long-term reservoir of ESBL *Enterobacteriaceae*. Colonizing ESBL *Enterobacteriaceae* were all identified as *E. coli* including strains that are unlikely to be pathogenic and strains carrying virulence genes associated with extraintestinal pathogenic *E. coli*. Finally, many strains carried multidrug resistance plasmids and we observed at least one participant where the same plasmid was observed in isolates with different phylogenetic backgrounds, consistent with a role for the human gut as a hotspot for antibiotic resistance gene exchange (74). Overall, our study suggests that the human gut may represent an important but under-recognized reservoir of ESBL genes and ESBL-carrying *E. coli and* highlights the relevance and importance of understanding the role of gut commensals in mediating the spread of antibiotic resistance.

## Materials and Methods

### Specimen collection, metadata collection and analysis

Recruitment and consenting of participants were performed by the Clinical and Translational Research Unit (CTRU) at the University of Georgia. Inclusion criteria included: the ability and willingness to answer an online survey regarding risk factors and to physically visit the CTRU to obtain and return the specimen collection kit, as well as age >18 years for adult participants. Adult participants with a child age ≤ 5 years in their household were invited to enroll the child in the study. Exclusion criteria included pregnancy and in-patient (overnight) hospitalization/health care in the last 12 months for reasons other than uncomplicated childbirth. Prospective participants who reported systemic (oral or intravenous) antibiotic use within the last 48 hours were asked to schedule specimen collection/drop-off for a later date, as were participants with active COVID-19 infections. Children who did not reside with participant parent for 5 days or more per week were also excluded. A signed consent was obtained from each participant or parent. To protect the confidentiality of personal data, all participants were assigned a unique, randomly generated identification number. All research activities involving human subject research were reviewed and approved by the Institutional Review Board (IRB) at University of Georgia, Athens.

Participants were provided a stool specimen container that was pre-labeled with their participant ID (Medline Industries, Cat. No. DYND36500) with a scheduled sample return date. Participants were asked to collect a stool sample using the provided collection kit and to complete an online questionnaire to collect demographic information as well as possible environmental risk factors for carriage of antibiotic resistant bacteria (see supplemental materials) as close in time as possible and no more than 12 hours before their scheduled return appointment. After collection, they were instructed to keep the stool specimen in a refrigerator or protected from heat until return to the facility. Upon return to the CTRU, stool specimens were stored at 4°C until processing. Upon transfer to the study team, stool specimens were subdivided for processing, typically within 1-3 hours of receipt (max. 24 hours). For culture work, 200 mg of stool specimens were diluted in 1 mL of 1X PBS and immediately processed as described below. For DNA extraction and molecular analyses, 200 mg sub-specimens were transferred to sterile cryovials and stored at −20°C until processing as described below.

Participant data was collected by online survey (Qualtrics) and matched to laboratory samples via alphanumeric participant IDs. Participant surveys consisted of a series of questions relating to each participant’s demographics, lifestyle, socioeconomic status, environmental exposures, preexisting medical conditions, and travel experience. All child surveys were completed by a designated parent or guardian. Upon completion of sample collection, all data were downloaded, matched to laboratory results, and cleaned for analysis using R statistical analysis software with ‘tidyverse’, ‘readxl’, ‘magrittr’, ‘reshape2’ and ‘rcompanion’. ESBL carriage frequencies and Fisher’s test with Monte-Carlo simulations were used to calculate p-values using ‘fisher.test’ function in R. P-adjusted values were calculated using the ‘p.adjust’ function with the Benjamini-Hochberg procedure for False Discovery Rate (FDR) (75, 76). Statistical significance was considered from p-values less than 0.05.

### Isolation, testing and confirmation of ESBL-E

200 mg of each stool specimen was diluted in 1 ml of 1X PBS. 100µl of the diluted aliquot was pre-enriched overnight in 5 ml tryptic soy broth (TSB) at 37°C. 10 µl of the enriched culture was then spread onto CHROMagar^TM^ ESBL plates and incubated at 37°C overnight. Well-isolated colonies from CHROMagar ESBL plates were re-streaked for isolation and purification on the same medium and incubated under the same conditions. Presumptive ESBL-producing colonies were re-streaked on blood agar, identified by MALDI-TOF MS, and confirmed by antibiotic resistant profiling using Sensititre panel ESB1F at the UGA Veterinary Diagnostics Laboratory. An isolate was considered ESBL-positive if it was resistant to Cefotaxime and Ceftazidime but susceptible when clavulanic acid was added to each. Other antibiotics tested with Sensititre included Ampicillin, Cefazolin, Cefepime, Cefoxitin, Cefpodoxime, Ceftriaxone, Cephalothin, Ciprofloxacin, Gentamicin, Imipenem, Meropenem and Piperacillin/Tazobactam. Confirmed ESBL-positive participants were requested to provide a second sample at least 90 days after the initial participation. 22 of 25 positive participants provided a second sample, the remaining 3 could not be contacted after multiple attempts. Second-visit samples were processed under the same conditions as the first samples.

### Whole-genome sequencing and analysis

Isolated colonies from ESBL-confirmed participants were used for whole-genome analysis. Genomic DNA of isolates was extracted using Omega Biotek Bacterial DNA Kit. Purified DNA was arrayed in 96-well plates, normalized, and run on agarose minigels for QC. Genomic libraries were created using an NEBNext Ultra II FS DNA Library Prep Kit with custom primers and protocols (77) and an Opentrons OT-2 robot. Libraries for all ESBL-confirmed isolates were sequenced on a NovaSeq S4 6000 to obtain PE150 reads. Trimming and quality filtering was performed via Trimmomatic v0.39.

For long-read sequencing, DNA extracts were normalized to 50 ng per 9 µl. Oxford Nanopore Technologies (ONT) libraries were prepared following manufacturer’s instructions for the rapid barcode 96 kit and cleaned with Ampure XP beads. A total of 75 µl of library was loaded onto an ONT MinION flow cell (9.3.1) for sequencing. Following sequencing, bases were called using super accuracy mode with Guppy v6.1.1.

*De novo* assembly was performed using Unicycler v0.4.7. using the short-reads first approach and default settings (78). A phylogenetic tree between ESBL-E genomes was constructed using maximum likelihood and core single nucleotide polymorphisms with Parsnp, with a randomly selected reference genome (AREA_ 490). Core genome single nucleotide polymorphisms (SNPs) between isolates from first and second samples were calculated using CSI Phylogeny (79, 80). AMRFinder v3.9.8 was used to identify antimicrobial resistance genes in assembled genomes using the Plus genes database (--plus) and *E. coli* as the reference organism (-O) to identify point mutations (81). Mobile genetic elements were identified using MobileElementFinder v1.0.3 with antimicrobial resistance genes annotated (82). Tools from the Center for Genomic Epidemiology (http://www.genomicepidemiology.org/services/) that were used to analyze the hybrid assemblies including: MLST v2.0.9 with configuration for *Escherichia coli* #1 and a minimum depth of 5X; SeroTypeFinder v2.0.1 with 85% threshold ID and 60% minimum length, FimTyper v1.0 with 95% threshold ID, 95% threshold identity and 60% minimum coverage and VirulenceFinder v2.0.3 for *E. coli* with 90% threshold ID and 60% minimum length (83–86). Assemblies were classified by phylogroups using the ClermonTyping web server (http://clermontyping.iame-research.center)(87).

### Microbial community characterization

Fecal *E*. *coli* were isolated without antibiotic selection from all stool samples using CHROMagar ECC. Up to 48 well-isolated colonies (if available) were re-streaked on LB plates for purity, then arrayed in 96-well plates to grow overnight with Mueller Hinton II broth media. 3 µl subsamples of the overnight culture were patched in Mueller Hinton II agar plates containing antibiotics at CLSI standards: Ampicillin (32 µg/ml), Ceftriaxone (32 µg/ml), Ciprofloxacin (4 µg/ml), Tetracycline (4 µg/ml) and Trimethoprim (8 µg/ml) to characterize the AR profile of the commensal *E. coli* community in the ESBL-E positive participants.

For 16S rRNA gene library sequencing, DNA was isolated from 200 mg of each stool sample with the Omega Biotek Stool DNA kit following the manufacturer protocol for pathogen detection (Omega Biotek, Norcross, GA, USA). The optional incubation with DS buffer and Proteinase K was performed to improve recovery of Gram-positive bacteria. Extracted DNA was eluted in 100µl of elution buffer from the kit heated at 65°C. DNA concentrations were measured with NanoDrop. Amplification of 16S rRNA gene was done using primers 515F and 806R as described previously (88), followed by library sequencing with Illumina MiSeq 250 x 250 bp at the Georgia Genomics Facility. Sequence analysis was performed using DADA2 (version 1.18) in R (version 4.0.2) including filtering, trimming, merge and taxonomy assignment with SILVA database, version 138 (89, 90). Data analyses were done in R using phyloseq, vegan, dplyr, tidyverse and DESeq packages with samples rarefied to a depth of 10,000 when needed (91, 92).

## Data availability

Whole-genome assemblies and 16S rDNA sequences have been submitted to NCBI under the BioProject: PRJNA894544.

## Acknowledgements

This work was supported by BAA contract no. 75D30120C09496 from the Centers for Disease Control and Prevention. Funders had no role in data collection and interpretation, or the decision to submit the publication. Participants recruitment and sample processing was performed with the help of the CTRU at University of Georgia, which is supported by the National Center for Advancing Translational Sciences of the National Institutes of Health under award number: UL1TR002378. The content is solely the responsibility of the authors and does not necessarily represent the official views of the National Institutes of Health.

Author roles based on CRediT taxonomy: Coralis Rodriguez Garcia, data curation, formal analysis, investigation, methodology, visualization, writing – original draft; William A. Norfolk, data curation, formal analysis, visualization; Amanda K. Howard, investigation, methodology, formal analysis; Amanda L. Glatter, investigation; Megan S. Beaudry, investigation; Nicholas A. Mallis, investigation; Michael Welton, conceptualization, methodology; Travis C. Glenn, conceptualization, methodology, supervision, project administration; Erin K. Lipp, conceptualization, methodology, supervision, project administration; Elizabeth A. Ottesen, conceptualization, funding acquisition, methodology, supervision, project administration, writing – review and editing.

We also thank Andreas Handel, Jonathan Frye, and Charlene Jackson for their advice regarding methodology and data analysis, as well as Marissa Howard, Kelly Petersen, Bryce Miller, Katie Dillon, Kirty Muthyala, Claire Pearson for their assistance in data collection.

