## Supplemental Materials for "Long-term gut colonization with ESBL-producing *Escherichia coli* in participants without known risk factors from the southeastern United States"

**Table of Content**

Tables:

Table S1 ------------------------------------------------------------ 2
Table S2 ------------------------------------------------------------ 6
Table S3 ------------------------------------------------------------ 7
Table S4 ------------------------------------------------------------ 9
Table S5 ------------------------------------------------------------ 12

Figures:

Figure S1 ------------------------------------------------------------ 14
Figure S2 ------------------------------------------------------------ 15
Figure S3 ------------------------------------------------------------ 16
Figure S4 ------------------------------------------------------------ 17
Figure S5 ------------------------------------------------------------ 18
Figure S6 ------------------------------------------------------------ 19
Figure S7 ------------------------------------------------------------ 20
Figure S8 ------------------------------------------------------------ 21
Figure S9 ------------------------------------------------------------ 21

**Table S1**: Observed and expected values for ESBL-E carriage on selected demographic information collected from the questionnaire filled by all adult participants during their first sample provided.

| Demographics or Risk Factors | N | ESBL-E Positive (N=24) | ESBL-E Negative (N=481) | p-value/ FDR p-adjusted |
|---|---|---|---|---|
| **Biological Sex** | | | | |
| Female | 349 | 13 (4%) | 336 (96%) | 0.12/0.79 |
| Male | 156 | 11 (7%) | 145 (93%) | |
| **Race** | | | | |
| Asian | 37 | 4 (11%) | 33 (89%) | 0.17/0.79 |
| Black or African American | 59 | 5 (8%) | 54 (92%) | |
| White | 377 | 15 (4%) | 362 (96%) | |
| **Age** | | | | |
| 18-29 | 234 | 13 (6%) | 221 (94%) | 0.17/0.79 |
| 30-39 | 84 | 2 (2%) | 82 (98%) | |
| 40-49 | 53 | 3 (6%) | 50 (94%) | |
| 50-59 | 49 | 2 (4%) | 47 (96%) | |
| 60-69 | 45 | 1 (2%) | 44 (98%) | |
| 70-79 | 22 | 2 (9%) | 20 (91%) | |
| 80-89 | 5 | 0 (0%) | 5 (100%) | |
| **Currently Employed** | | | | |
| Yes | 340 | 12 (4%) | 328 (96%) | 0.08/0.79 |
| No | 165 | 12 (7%) | 153 (93%) | |
| **Ethnicity** | | | | |
| Hispanic | 42 | 1 (2%) | 41 (98%) | 0.84/1 |
| Non-Hispanic | 450 | 23 (5%) | 427 (95%) | |
| **Highest Schooling** | | | | |
| Less Than High School | 5 | 0 (0%) | 5 (100%) | 0.14/0.79 |
| High School/GED | 54 | 2 (4%) | 52 (96%) | |
| Some College | 138 | 11 (8%) | 127 (92%) | |
| Associate degree | 39 | 3 (8%) | 36 (92%) | |
| Bachelor's Degree | 149 | 5 (3%) | 144 (97%) | |
| Master's Degree | 97 | 1 (1%) | 96 (99%) | |
| Ph.D., MD, or JD | 23 | 2 (9%) | 21 (91%) | |
| **Annual Household Income** | | | | |
| Less than $20,000 | 96 | 5 (5%) | 91 (95%) | 0.45/0.79 |
| $20,000 to $34,999 | 72 | 6 (8%) | 66 (92%) | |
| $35,000 to $49,999 | 51 | 3 (6%) | 48 (94%) | |
| $50,000 to $74,999 | 84 | 2 (2%) | 82 (98%) | |

| | | | | |
|---|---|---|---|---|
| $75,000 to $99,999 | 63 | 4 (6%) | 59 (94%) | |
| Over $100,000 | 132 | 4 (3%) | 128 (97%) | |
| **Residence Type** | | | | |
| Single Family Home | 292 | 11 (4%) | 281 (96%) | 0.25/0.79 |
| Apartment/Multi Family | 209 | 13 (6%) | 196 (94%) | |
| Homeless | 1 | 0 (0%) | 1 (100%) | |
| **Rural, Urban or Suburban** | | | | |
| Urban | 130 | 5 (4%) | 125 (96%) | 0.68/0.99 |
| Rural | 91 | 3 (3%) | 88 (97%) | |
| Suburban | 282 | 16 (6%) | 266 (94%) | |
| **Primary Water Source** | | | | |
| Town/Municipal Water | 441 | 21 (5%) | 420 (95%) | 0.18/0.79 |
| Private Well | 49 | 1 (2%) | 48 (98%) | |
| Other | 15 | 2 (13%) | 13 (87%) | |
| **Exposure to animals at home, farm, or animal care facility**[1] | | | | |
| Dogs and/or Cats | 251 | 9 (4%) | 242 (96%) | 0.42/0.79 |
| Other than dog or cat | 16 | 0 (0%) | 16 (100%) | |
| Multiple Types | 24 | 1 (24%) | 23 (96%) | |
| None | 214 | 14 (7%) | 200 (93%) | |
| **Regular livestock exposure** | | | | |
| Yes | 36 | 0 (0%) | 36 (100%) | 0.40/0.79 |
| No | 468 | 23 (5%) | 445 (95%) | |
| **Number of risk environments exposed to**[2] | | | | |
| 1 | 85 | 3 (4%) | 82 (96%) | 0.89/1 |
| 2 | 44 | 0 (0%) | 44 (100%) | |
| 3 | 20 | 2 (10%) | 18 (90%) | |
| 4 | 7 | 0 (0%) | 7 (100%) | |
| 5 | 6 | 0 (0%) | 6 (100%) | |
| 6 | 1 | 0 (0%) | 1 (100%) | |
| 7 | 1 | 0 (0%) | 1 (100%) | |
| None | 333 | 17 (5%) | 316 (95%) | |
| **Exposure to healthcare facilities (occupational or visitor)** | | | | |
| Yes | 81 | 4 (5%) | 77 (95%) | 0.78/0.99 |
| No | 412 | 19 (5%) | 393 (95%) | |
| **Food poisoning in last 30 days** | | | | |
| Yes | 5 | 0 (0%) | 5 (100%) | 1/1 |
| No | 500 | 24 (5%) | 476 (95%) | |
| **Gastrointestinal conditions or symptoms in the past 1 month** | | | | |
| Yes | 177 | 8 (5%) | 169 (95%) | 1/1 |

| | | | | |
|---|---|---|---|---|
| No | 327 | 16 (5%) | 311 (95%) | |
| **Multiple antibiotics taken this year** | | | | |
| Yes | 40 | 3 (8%) | 37 (92%) | 0.43/0.79 |
| No | 465 | 21 (5%) | 444 (95%) | |
| **Antibiotics use in the past 1 month (oral, topical, or intravenous)** | | | | |
| Yes | 47 | 2 (4%) | 45 (96%) | 1/1 |
| No | 454 | 22 (5%) | 432 (95%) | |
| **Regular medications or supplements taken in last 30 days** | | | | |
| Yes | 363 | 14 (3%) | 348 (96%) | 0/0.01 |
| No | 40 | 9 (2%) | 31 (40%) | |
| **Urinary tract infection in the past 1 month (self-diagnosed or diagnosed by doctor)** | | | | |
| Yes | 13 | 1 (8%) | 12 (92%) | 0.47/0/79 |
| No | 492 | 23 (5%) | 469 (95%) | |
| **International travel (past year)** | | | | |
| Yes | 31 | 1 (3%) | 30 (97%) | 1/1 |
| No | 471 | 23 (5%) | 448 (95%) | |
| **Lived internationally in the last 5 years** | | | | |
| Yes | 37 | 4 (11%) | 33 (89%) | 0.09/0.79 |
| No | 463 | 20 (4%) | 443 (96%) | |
| **Exposure to treated recreational water in the past month** | | | | |
| Yes | 127 | 4 (3%) | 123 (97%) | 0.47/0.79 |
| No | 371 | 20 (5%) | 351 (95%) | |
| **Exposure to untreated recreational water in the past month** | | | | |
| Yes | 107 | 3 (3%) | 104 (97%) | 0.44/0.79 |
| No | 391 | 21 (5%) | 370 (95%) | |
| **Additional house members** | | | | |
| 0 | 57 | 3 (5%) | 54 (95%) | 0.74/0.99 |
| >1 | 447 | 21 (5%) | 426 (95%) | |
| **Additional house members younger than 5 years old** | | | | |
| 0 | 400 | 19 (5%) | 381 (95%) | 1/1 |
| >1 | 92 | 4 (4%) | 88 (96%) | |
| **Eat poultry in the last week** | | | | |
| Yes | 421 | 21 (5%) | 400 (95%) | 0.78/0.99 |
| No | 83 | 3 (4%) | 80 (96%) | |
| **Eat pork or beef in the last week** | | | | |
| Yes | 385 | 16 (4%) | 369 (96%) | 0.32/0.79 |
| No | 119 | 8 (7%) | 111 (93%) | |
| **Eat fish or shellfish in the last week** | | | | |
| Yes | 254 | 10 (4%) | 244 (96%) | 0.41/0.79 |

| | | | | |
|---|---|---|---|---|
| No | 248 | 14 (6%) | 234 (94%) | |
| **Eat dairy in the last week** | | | | |
| Yes | 473 | 24 (5%) | 449 (95%) | 0.39/0.79 |
| No | 32 | 0 (0%) | 32 (100%) | |
| **Eat raw fruit or vegetables in the last week** | | | | |
| Yes | 439 | 19 (4%) | 420 (96%) | 0.22/0.79 |
| No | 65 | 5 (8%) | 60 (92%) | |

[1]Animals reported in 'Other than dog or cat' include reptiles, birds, rodents/small mammals. If participant reported dog and/or cat in addition to other types, then it was classified in 'Multiple.' [2]Questionnaire offered multiple choices as environmental risk exposure including animal waste, human waste, companion animals, pesticides/herbicides, childcare facilities, K-12 schools, raw meat/poultry, poultry, livestock, veterinary facilities and correctional facilities, data reported includes the number of environments selected.

**Table S2**: Demographic distribution from our study compared to Athens-Clarke County, the state of Georgia and over the U.S. based on data collected from census.gov (2021).

| **Demographics** | **Distribution of adults on this study (N=505)** | **Distribution in Athens Clarke County** | **Distribution in Georgia** | **Distribution in the U.S.A*** |
|---|---|---|---|---|
| *Asian* | 7.3% | 4.0% | 4.8% | 5.8% |
| *Black or African American* | 11.7% | 27.8% | 33.1% | 12.1% |
| *White* | 74.6% | 60.5% | 59.0% | 61.2% |
| *Mixed* | 1.8% | 4.4% | 2.4% | 12.6% |
| *Other* | 2.4% | - | - | - |
| *Hispanic* | 8.3% | | | 18.8% |
| *Non-Hispanic* | 89.1% | | | 81.2% |
| *Identified as Female* | 69.1% | | | 50.5% |
| *Identified as Male* | 30.8% | | | 49.5% |

*From American Community Survey 2-21: ACS-1 year estimates data profiles

**Table S3: Genome quality assessment using CheckM lineage workflow**

| Isolate | Genome Size (bp) | Completeness (%) | Contamination (%) | Total of N50 contigs | Longest contig (bp) | Mean contig length (bp) |
|---|---|---|---|---|---|---|
| AREA_482 | 4,981,979 | 99.07 | 0.52 | 157,133 | 398,159 | 25,418 |
| AREA_483 | 5,438,138 | 99.97 | 0.07 | 5,106,887 | 5,106,887 | 543,814 |
| AREA_484 | 5,299,331 | 99.97 | 0.45 | 5,126,651 | 5,126,651 | 1,059,866 |
| AREA_485 | 5,461,929 | 99.97 | 0.33 | 5,310,577 | 5,310,577 | 1,820,643 |
| AREA_486 | 5,172,013 | 99.97 | 0.39 | 5,101,202 | 5,101,202 | 1,724,004 |
| AREA_487 | 5,270,828 | 99.97 | 0.33 | 5,189,749 | 5,189,749 | 1,756,943 |
| AREA_488 | 5,225,697 | 99.97 | 0.36 | 1,341,054 | 1,703,105 | 348,380 |
| AREA_489 | 5,235,109 | 99.97 | 0.33 | 3,263,269 | 3,263,269 | 193,893 |
| AREA_490 | 5,444,513 | 99.97 | 0.60 | 3,725,720 | 3,725,720 | 494,956 |
| AREA_491 | 5,303,945 | 99.82 | 0.35 | 678,206 | 728,890 | 88,399 |
| AREA_492 | 5,154,401 | 99.67 | 0.72 | 4,934,235 | 4,934,235 | 644,300 |
| AREA_493 | 5,141,066 | 99.97 | 0.39 | 4,722,299 | 4,722,299 | 1,028,213 |
| AREA_494 | 5,373,309 | 99.97 | 0.33 | 5,164,941 | 5,164,941 | 1,791,103 |
| AREA_495 | 5,260,164 | 99.07 | 0.49 | 822,129 | 2,523,629 | 328,760 |
| AREA_496 | 5,335,487 | 99.97 | 0.43 | 5,183,188 | 5,183,188 | 1,778,496 |
| AREA_497 | 4,880,542 | 99.93 | 0.10 | 4,699,031 | 4,699,031 | 1,626,847 |
| AREA_498 | 5,427,885 | 99.97 | 1.33 | 4,679,123 | 4,679,123 | 904,648 |
| AREA_499 | 5,203,150 | 99.97 | 0.39 | 5,203,150 | 5,203,150 | 5,203,150 |
| AREA_500 | 5,327,239 | 99.97 | 0.36 | 5,108,113 | 5,108,113 | 197,305 |
| AREA_501 | 5,210,053 | 99.37 | 0.33 | 687,543 | 2,330,037 | 89,829 |
| AREA_502 | 5,141,719 | 99.97 | 0.33 | 5,039,263 | 5,039,263 | 1,713,906 |
| AREA_503 | 5,274,630 | 99.97 | 0.33 | 5,150,049 | 5,150,049 | 1,758,210 |

| | | | | | | |
|---|---|---|---|---|---|---|
| AREA_504 | 5,275,050 | 99.97 | 0.45 | 5,081,105 | 5,081,105 | 1,318,763 |
| AREA_505 | 5,039,223 | 99.97 | 0.33 | 4,916,587 | 4,916,587 | 2,519,612 |
| AREA_C483 | 5,074,575 | 99.93 | 1.24 | 5,074,575 | 5,074,575 | 5,074,575 |
| AREA_R482 | 5,103,194 | 99.67 | 0.37 | 323,367 | 522,390 | 60,038 |
| AREA_R483 | 5,436,943 | 99.91 | 0.17 | 1,148,558 | 1,580,857 | 84,952 |
| AREA_R485 | 5,465,567 | 99.97 | 0.48 | 5,315,488 | 5,315,488 | 2,732,784 |
| AREA_R488 | 5,193,622 | 99.97 | 0.36 | 5,059,100 | 5,059,100 | 78,691 |
| AREA_R489 | 5,224,086 | 99.97 | 0.33 | 3,260,324 | 3,260,324 | 200,926 |
| AREA_R490 | 5,356,116 | 99.07 | 0.51 | 691,421 | 2,118,936 | 198,375 |
| AREA_R491 | 5,358,613 | 99.97 | 0.29 | 5,236,147 | 5,236,147 | 1,071,723 |
| AREA_R493 | 5,112,865 | 99.97 | 0.39 | 1,013,536 | 2,468,508 | 340,858 |
| AREA_R497 | 5,047,689 | 99.97 | 0.06 | 3,122,400 | 3,122,400 | 630,961 |
| AREA_R500 | 5,279,145 | 99.79 | 0.51 | 4,847,616 | 4,847,616 | 55,570 |
| AREA_R501 | 5,229,413 | 99.97 | 0.33 | 3,849,269 | 3,849,269 | 116,209 |
| AREA_R503 | 5,274,177 | 99.97 | 0.33 | 3,889,283 | 3,889,283 | 1,758,059 |
| AREA_R504 | 5,286,355 | 99.97 | 0.45 | 5,082,024 | 5,082,024 | 881,059 |
| AREA_R505 | 5,040,783 | 99.97 | 0.33 | 4,918,147 | 4,918,147 | 2,520,392 |

**Table S4**: Assembly results by isolate performed by Unicycler and visualized in Bandage. Some plasmids were assembled and circularized by Unicycler but not identified in PlasmidFinder. Complete and incomplete assemblies are available in NCBI.

| Isolate | Scaffold length | Scaffold status | Scaffold ID | NCBI BioSample Accession Number |
|---|---|---|---|---|
| AREA_482 | 1. 4,901,008<br>2. 93,130 | 1. Incomplete<br>2. Incomplete | 1. Chromosome<br>2. Plasmid | SAMN33912483 |
| AREA_R482 | 1. 4,900,803<br>2. 111,841<br>3. 94,195 | 1. Incomplete<br>2. Incomplete<br>3. Incomplete | 1. Chromosome<br>2. Plasmid<br>3. Plasmid | SAMN33912484 |
| AREA_483 | 1. 5,106,888<br>2. 85,875<br>3. 83,837<br>4. 95,437<br>5. 59,126<br>6. 6,989 | 1. Complete<br>2. Complete<br>3. Complete<br>4. Incomplete<br>5. Complete<br>6. Complete | 1. Chromosome<br>2. Plasmid<br>3. Plasmid<br>4. Plasmid<br>5. Plasmid<br>6. Plasmid | SAMN33912485 |
| AREA_R483 | 1. 3,960,029<br>2. 1,148,558<br>3. 95,437<br>4. 85,876<br>5. 83,837<br>6. 59,124<br>7. 6,989 | 1. Incomplete<br>2. Incomplete<br>3. Complete<br>4. Complete<br>5. Complete<br>6. Complete<br>7. Complete | 1. Chromosome<br>2. Chromosome<br>3. Plasmid<br>4. Plasmid<br>5. Plasmid<br>6. Plasmid<br>7. Plasmid | SAMN33912486 |
| AREA_484 | 1. 5,126,947<br>2. 93,541<br>3. 70,016<br>4. 8,828 | 1. Incomplete<br>2. Complete<br>3. Complete<br>4. Complete | 1. Chromosome<br>2. Plasmid<br>3. Plasmid<br>4. Plasmid | SAMN33912487 |
| AREA_485 | 1. 5,310,580<br>2. 149,251<br>3. 2,101 | 1. Incomplete<br>2. Complete<br>3. Complete | 1. Chromosome<br>2. Plasmid<br>3. Plasmid | SAMN33912488 |
| AREA_R485 | 1. 5,315,468<br>2. 150,079 | 1. Complete<br>2. Complete | 1. Chromosome<br>2. Plasmid | SAMN33912489 |
| AREA_486 | 1. 5,101,199<br>2. 65,601<br>3. 5,210 | 1. Complete<br>2. Complete<br>3. Complete | 1. Chromosome<br>2. Plasmid<br>3. Plasmid | SAMN33912490 |
| AREA_487 | 1. 5,189,868<br>2. 79,530<br>3. 1,549 | 1. Complete<br>2. Complete<br>3. Complete | 1. Chromosome<br>2. Plasmid<br>3. Plasmid | SAMN33912491 |
| AREA_488 | 1. 5,058,538<br>2. 96,059<br>3. 68,388<br>4. 3,257 | 1. Incomplete<br>2. Incomplete<br>3. Complete<br>4. Complete | 1. Chromosome<br>2. Plasmid<br>3. Plasmid<br>4. Plasmid | SAMN33912492 |
| AREA_R488 | 1. 5,059,104<br>2. 135,828<br>3. 3,257 | 1. Complete<br>2. Incomplete<br>3. Complete | 1. Chromosome<br>2. Plasmid<br>3. Plasmid | SAMN33912493 |
| AREA_489 | 1. 5,091,059<br>2. 107,575<br>3. 30,657<br>4. 7,939 | 1. Incomplete<br>2. Complete<br>3. Complete<br>4. Complete | 1. Chromosome<br>2. Plasmid<br>3. Plasmid<br>4. Plasmid | SAMN33912494 |
| AREA_R489 | 1. 5,088,296 | 1. Incomplete | 1. Chromosome | SAMN33912495 |

| | | | | |
|---|---|---|---|---|
| | 2. 107,566<br>3. 30,649 | 2. Complete<br>3. Complete | 2. Plasmid<br>3. Plasmid | |
| AREA_490 | 1. 5,257,305<br>2. 123,216<br>3. 47,546<br>4. 7,939<br>5. 5,269<br>6. 3,256 | 1. Incomplete<br>2. Complete<br>3. Complete<br>4. Complete<br>5. Complete<br>6. Complete | 1. Chromosome<br>2. Plasmid<br>3. Plasmid<br>4. Plasmid<br>5. Plasmid<br>6. Plasmid | SAMN33912496 |
| AREA_R490 | 1. 2,119,052<br>2. 1,403,314<br>3. 1,653,205<br>4. 123,216<br>5. 47,546<br>6. 5,269<br>7. 4,063<br>8. 1,115 | 1. Incomplete<br>2. Incomplete<br>3. Incomplete<br>4. Complete<br>5. Complete<br>6. Complete<br>7. Complete<br>8. Incomplete | 1. Chromosome<br>2. Chromosome<br>3. Chromosome<br>4. Plasmid<br>5. Plasmid<br>6. Plasmid<br>7. Plasmid<br>8. Plasmid | SAMN33912497 |
| AREA_491 | 1. 5,186,701<br>2. 116,786<br>3. 4,076 | 1. Incomplete<br>2. Complete<br>3. Complete | 1. Chromosome<br>2. Plasmid<br>3. Plasmid | SAMN33912498 |
| AREA_R491 | 1. 5,242,716<br>2. 115,977 | 1. Incomplete<br>2. Complete | 1. Chromosome<br>2. Plasmid | SAMN33912499 |
| AREA_492 | 1. 4,934,455<br>2. 123,608<br>3. 71,888<br>4. 7,939<br>5. 5,430<br>6. 5,166<br>7. 4,073<br>8. 2,077 | 1. Complete<br>2. Complete<br>3. Complete<br>4. Complete<br>5. Complete<br>6. Complete<br>7. Complete<br>8. Complete | 1. Chromosome<br>2. Plasmid<br>3. Plasmid<br>4. Plasmid<br>5. Plasmid<br>6. Plasmid<br>7. Plasmid<br>8. Plasmid | SAMN33912500 |
| AREA_493 | 1. 4,961,184<br>2. 109,539<br>3. 70,343 | 1. Incomplete<br>2. Complete<br>3. Complete | 1. Chromosome<br>2. Plasmid<br>3. Plasmid | SAMN33912501 |
| AREA_R493 | 1. 5,009,157<br>2. 70,343<br>3. 34,615 | 1. Incomplete<br>2. Complete<br>3. Complete | 1. Chromosome<br>2. Plasmid<br>3. Plasmid | SAMN33912502 |
| AREA_494 | 1. 5,164,944<br>2. 110,786<br>3. 97,582 | 1. Complete<br>2. Complete<br>3. Complete | 1. Chromosome<br>2. Plasmid<br>3. Plasmid | SAMN33912503 |
| AREA_495 | 1. 4,852,083<br>2. 309,561<br>3. 76,447<br>4. 22,159 | 1. Incomplete<br>2. Complete<br>3. Complete<br>4. Incomplete | 1. Chromosome<br>2. Plasmid<br>3. Plasmid<br>4. Plasmid | SAMN33912504 |
| AREA_496 | 1. 5,238,477<br>2. 96,998 | 1. Incomplete<br>2. Complete | 1. Chromosome<br>2. Plasmid | SAMN33912505 |
| AREA_497 | 1. 4,699,025<br>2. 92,616<br>3. 88,895 | 1. Complete<br>2. Complete<br>3. Complete | 1. Chromosome<br>2. Plasmid<br>3. Plasmid | SAMN33912506 |
| AREA_497R | 1. 4,824,639<br>2. 88,895<br>3. 66,868 | 1. Incomplete<br>2. Complete<br>3. Complete | 1. Chromosome<br>2. Plasmid<br>3. Plasmid | SAMN33912507 |

| | | | | |
|---|---|---|---|---|
| | 4. 62,520<br>5. 5,167 | 4. Complete<br>5. Complete | 4. Plasmid<br>5. Plasmid | |
| AREA_498 | 1. 4,975,257<br>2. 211,174<br>3. 129,755<br>4. 111,678 | 1. Incomplete<br>2. Complete<br>3. Complete<br>4. Complete | 1. Chromosome<br>2. Plasmid<br>3. Plasmid<br>4. Plasmid | SAMN33912508 |
| AREA_499 | 1. 5,203,158 | 1. Complete | 1. Chromosome | SAMN33912509 |
| AREA_500 | 1. 5,108,101<br>2. 143,170<br>3. 68,037<br>4. 5,165<br>5. 3,257 | 1. Complete<br>2. Complete<br>3. Incomplete<br>4. Complete<br>5. Complete | 1. Chromosome<br>2. Plasmid<br>3. Chromosome<br>4. Plasmid<br>5. Plasmid | SAMN33912510 |
| AREA_R500 | 1. 4,988,230<br>2. 105,899<br>3. 185,370<br>4. 5,165 | 1. Incomplete<br>2. Incomplete<br>3. Incomplete<br>4. Complete | 1. Chromosome<br>2. Plasmid<br>3. Plasmid<br>4. Plasmid | SAMN33912511 |
| AREA_501 | 1. 5,003,752<br>2. 120,161<br>3. 86,612<br>4. 2,112 | 1. Incomplete<br>2. Complete<br>3. Complete<br>4. Complete | 1. Chromosome<br>2. Plasmid<br>3. Plasmid<br>4. Plasmid | SAMN33912512 |
| AREA_R501 | 1. 3,849,274<br>2. 1,176,107<br>3. 120,235<br>4. 86,643 | 1. Incomplete<br>2. Incomplete<br>3. Complete<br>4. Complete | 1. Chromosome<br>2. Chromosome<br>3. Plasmid<br>4. Plasmid | SAMN33912513 |
| AREA_502 | 1. 5,039,267<br>2. 97,288<br>3. 5,167 | 1. Complete<br>2. Complete<br>3. Complete | 1. Chromosome<br>2. Plasmid<br>3. Plasmid | SAMN33912514 |
| AREA_503 | 1. 5,171,793<br>2. 102,829 | 1. Incomplete<br>2. Complete | 1. Chromosome<br>2. Plasmid | SAMN33912515 |
| AREA_R503 | 1. 5,172,055<br>2. 102,819 | 1. Incomplete<br>2. Complete | 1. Chromosome<br>2. Plasmid | SAMN33912516 |
| AREA_504 | 1. 5,081,104<br>2. 114,903<br>3. 73,392<br>4. 5,631 | 1. Complete<br>2. Complete<br>3. Complete<br>4. Complete | 1. Chromosome<br>2. Plasmid<br>3. Plasmid<br>4. Plasmid | SAMN33912517 |
| AREA_R504 | 1. 5,082,014<br>2. 114,914<br>3. 73,403<br>4. 5,631<br>5. 5,214<br>6. 5,165 | 1. Complete<br>2. Complete<br>3. Complete<br>4. Complete<br>5. Complete<br>6. Complete | 1. Chromosome<br>2. Plasmid<br>3. Plasmid<br>4. Plasmid<br>5. Plasmid<br>6. Plasmid | SAMN33912518 |
| AREA_505 | 1. 4,916,592<br>2. 122,636 | 1. Complete<br>2. Complete | 1. Chromosome<br>2. Plasmid | SAMN33912519 |
| AREA_R505 | 1. 4,918,152<br>2. 122,636 | 1. Complete<br>2. Complete | 1. Chromosome<br>2. Plasmid | SAMN33912520 |
| AREA_C483 | 1. 5,074,572 | 1. Complete | 1. Chromosome | SAMN33912521 |

**Table S5: Assembled plasmids carried by each isolate identified by PlasmidFinder with encoded antibiotic genes identified by AMRFinderPlus and encoded virulence genes identified by VirulenceFinder**. Plasmid number in each isolate matches the numbers in panels C and D of Figure 1 and Figure 2.

| *Isolate* | Plasmid Type | Size (bp) | Antibiotic Resistance Genes | Virulence Genes |
|---|---|---|---|---|
| *497R* | 1. IncI1<br>2. IncFII<br>3. IncFII<br>4. Col156 | 1. 88,895<br>2. 66,870<br>3. 62,521<br>4. 5,167 | 1. *bla*$_{CTX-M-1}$ | 1. *cia*<br>2. *mcbA, traJ, traT* |
| *491* | 1. IncF<br>2. IncF | 1. 116,786<br>2. 4,076 | | |
| *498* | 1. IncFII<br>2. IncFIB<br>3. IncFIB (Phage) | 1. 211,173<br>2. 129,758<br>3. 111,678 | 1. –<br>2. *bla*$_{CTX-M-27}$ | 1. *aap, eatA, faeF, traT*<br>2. *traJ, traT* |
| *499* | - | - | | |
| *497* | 1. IncF<br>2. IncI1 | 1. 92,616<br>2. 88,895 | 1. *tetA, tetR*<br>2. *bla*$_{CTX-M-1}$ | 1. *anr, ompT*<br>2. *cia* |
| *495* | 1. IncF<br>2. IncF | 1. 309,561<br>2. 76,447 | 1. *bla*$_{CTX-M-15}$, *bla*$_{OXA-1}$, *bla*$_{TEM-1}$, *aph(3")-Ib, aph(6)-Id, aac(6')-Ib-cr5, catB3, floR, qacEdelta1, qnrS1, sul1, sul2, terB, terC, terD, terE, tet(A), dfrA1* | 1. *terC*<br>2. *anr, traJ, traT* |
| *483* | 1. IncFII<br>2. IncB/O/Z/K<br>3. IncI2 | 1. 83,837<br>2. 85,878<br>3. 59,126 | 1. *bla*$_{CTX-M-15}$, *qnrS1* | 1. *traT*<br>2. ireA |
| *C483* | - | - | | |
| *490* | 1. IncF<br>2. ColRNAI<br>3. Col440I | 1. 123,216<br>2. 7,939<br>3. 3,256 | 1. *tet(B)* | 1, *anr, iucC, iutA, sitA, traT* |
| *492* | 1. IncF<br>2. IncFII<br>3. ColRNAI<br>4. Col440II<br>5. Col156<br>6. Col (BS512) | 1. 123,593<br>2. 71,888<br>3. 7,939<br>4. 5,430<br>5. 5,166<br>6. 2,077 | 1. *bla*$_{TEM-1}$, *aph(3")-Ib, aph(6)-Id, sul2, tet(B)*<br>2. *bla*$_{TEM-1}$ | 1. *anr, iucC, iutA, sitA, traT*<br>2. *mcbA, traJ, traT* |
| *504* | 1. IncF<br>2. IncFII<br>3. Col156 | 1. 114,922<br>2. 73,392<br>3. 5,631 | 1. *bla*$_{CTX-M-27}$, *aadA5, aph(3")-Ib, aph(6)-Id, erm(B), mph(A), qacEdelta1, sul1, sul2, tet(A), dfrA17* | 1. *senB*<br>2. *fyuA, traJ* |
| *486* | 1. IncF<br>2. Col156 | 1. 65,601<br>2. 5,210 | 1. *bla*$_{CTX-M-27}$ | 1. *senB* |
| *482* | 1. IncF<br>2. Col156<br>3. IncY | 1. 74,522<br>2. 10,068<br>3. 88,765 | | 1. *traJ, traT*<br>2. *senB*<br>3. - |
| *505* | 1. IncF | 1. 122,636 | 1. *bla*$_{CTX-M-27}$, *aadA5, aph(3")-Ib, aph(6)-Id,* | 1. *anr, senB, traT* |

| | | | | |
|---|---|---|---|---|
| | | | *mph(A), qacEdelta1, sul1, sul2, tet(A), dfrA17* | |
| *493* | 1. IncF<br>2. IncF | 1. 109,542<br>2. 70,343 | 1. *bla*$_{CTX\text{-}M\text{-}27}$, *aadA5, aph(3")-Ib, aph(6)-Id, mph(A), qacEdelta1, sul1, sul2, tet(A), dfrA17* | 1. *anr, senB, traT*<br>2. *traT* |
| *496* | 1. IncF | 1. 96,998 | 1. *bla*$_{TEM\text{-}1}$, *aph(3")-Ib, aph(6)-Id, qnrS1, sul2, dfrA14* | 1. *anr, traJ, traT* |
| *503* | 1. IncF | 1. 102,829 | 1. *tet(A)* | 1. *iutA, senB* |
| *485* | 1. IncF<br>2. Col (BS512) | 1. 149,251<br>2. 2,101 | | 1. *anr, iutA, senB, traJ, traT* |
| *487* | 1. IncF<br>2. Col (MG828) | 1. 79,530<br>2. 1,549 | 1. *aph(3")-Ib, aph(6)-Id, sul2, tet(A)* | 1. *anr, senB* |
| *488* | 1. IncFI<br>2. IncFII<br>3. Col440I | 1. 96,050<br>2. 68,388<br>3. 3,257 | 1. *bla*$_{TEM\text{-}1}$, *aadA5, aph(3")-Ib, aph(6)-Id, mph(A), qacEdelta1, sul1, sul2, tet(A), dfrA17* | 1. *traJ, traT*<br>2. *mcbA, traT* |
| *500* | 1. IncF<br>2. Col156<br>3. Col440I | 1. 143,188<br>2. 5,165<br>3. 3,257 | 1. *bla*$_{TEM\text{-}1}$, *aac(3)-IId, aadA5, aph(3")-Ib, aph(6)-Id, mph(A), qacEdelta1, sul1, sul2, tet(A), dfrA17* | 1. *senB, traJ, traT* |
| *484* | 1. IncF<br>2. IncFII<br>3. IncQ1 | 1. 93,541<br>2. 70,016<br>3. 8,828 | 1. *bla*$_{TEM\text{-}1}$, *aac(3)-IId, aph(3")-Ib, aph(6)-Id, sul2, tet(B), dfrA17* | 1. *anr*<br>2. *traT* |
| *494* | 1. IncF<br>2. IncY | 1. 110,786<br>2. 97,582 | 1. *bla*$_{TEM\text{-}1}$ | 1. *anr, senB, traT* |
| *489* | 1. IncF<br>2. ColRNAI | 1. 107,575<br>2. 7,939 | 1. *bla*$_{CTX\text{-}M\text{-}27}$ | 1. *anr, senB, traT* |
| *501* | 1. IncF<br>2. IncI1<br>3. Col (BS512) | 1. 120,161<br>2. 86,606<br>3. 2,112 | 1. *bla*$_{CTX\text{-}M\text{-}27}$ | 1. *anr, senB, traT*<br>2. *cia* |
| *502* | 1. IncF<br>2. Col156 | 1. 97,289<br>2. 5,167 | 1. *bla*$_{CTX\text{-}M\text{-}27}$ | 1. *anr, senB* |

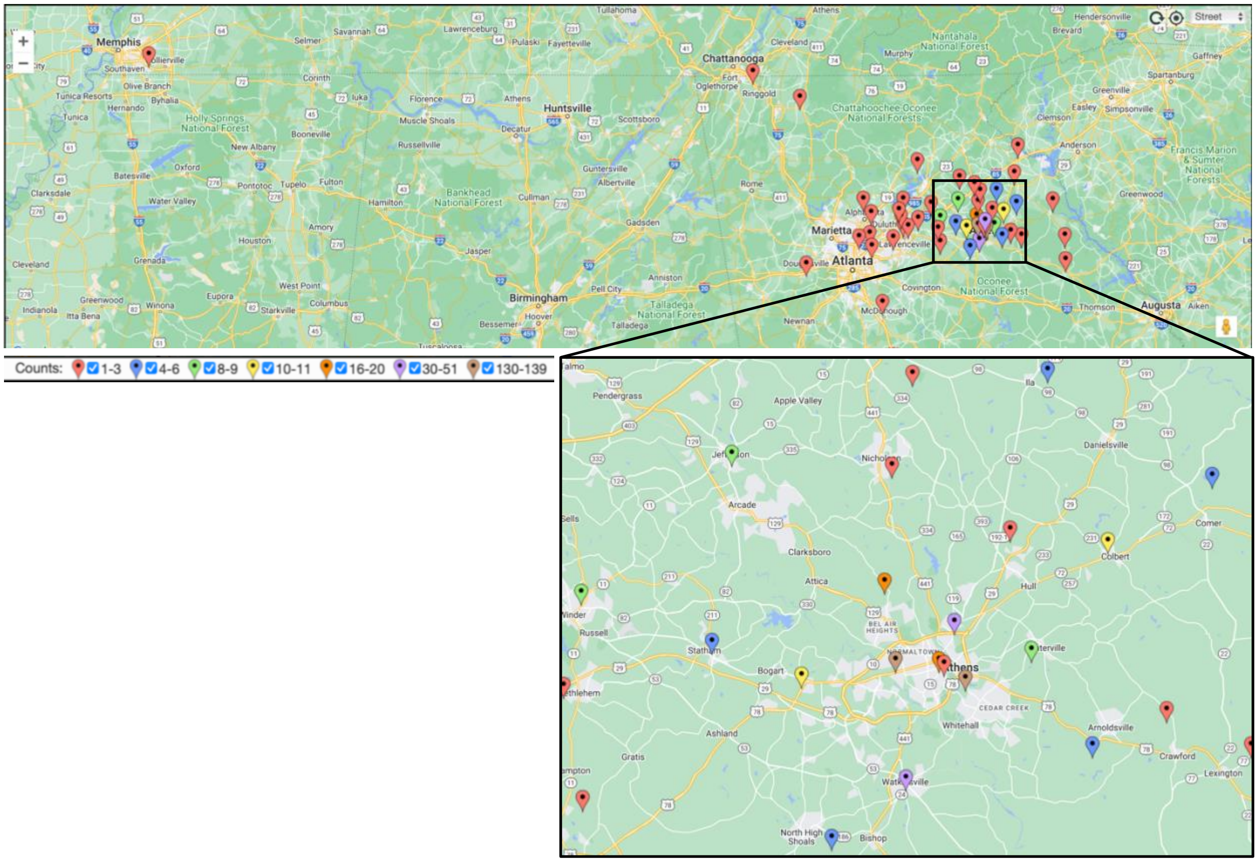


**Figure S1: Map of residence zip codes reported by adult participants**. The number of participants in each zip code is represented by color groups. Zip codes with more than four participants are limited to the Athens, GA area and vicinity. Map generated by EasyMapMaker.com.

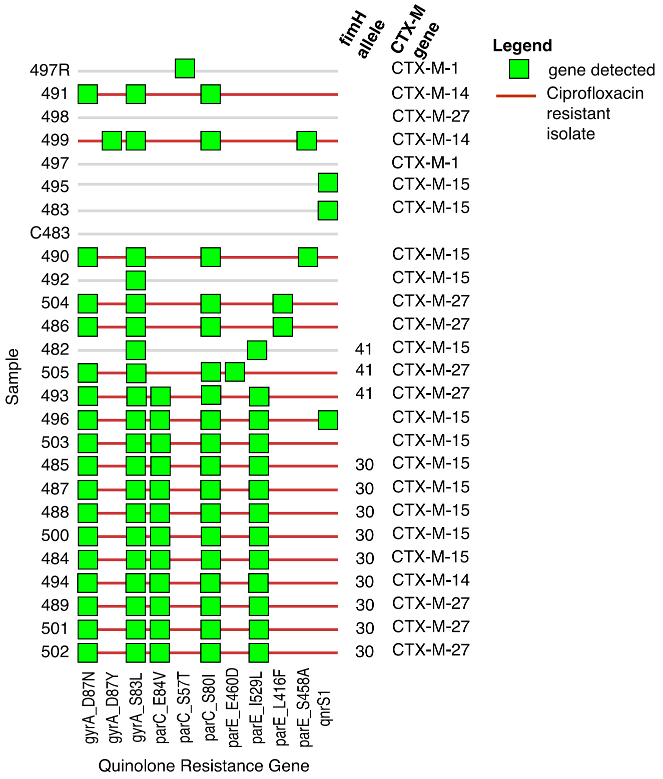


**Figure S2: Quinolone resistance phenotype and genotype of ESBL-E isolates**. Antimicrobial resistance genes from the quinolone class identified by AMRFinder on ESBL-E isolates are shown in green boxes. Samples that exhibited a Ciprofloxacin resistant phenotype are highlighted by red lines. For ESBL-E classified as ST131, their *fimH* allele is indicated, as well as their $bla_{CTX-M}$ allele if present for comparison of ST131 subclades.

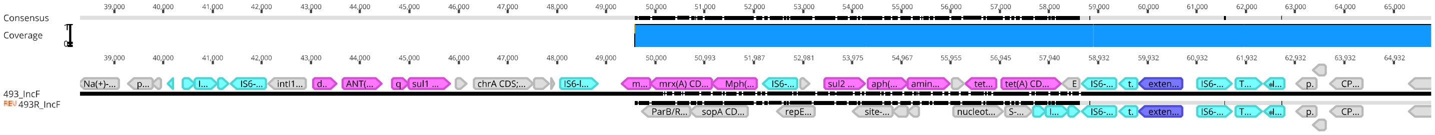


**Figure S3: Partial alignment of IncF plasmid in isolates 493 and 493R**. Alignment region showing insertion sequences (light blue) present in isolate 493 but missing in 493R, encoding multiple antibiotic resistance genes (pink). Beta-lactamase CTX-M-27 gene is shown in purple, also surrounded by IS6 sequences.

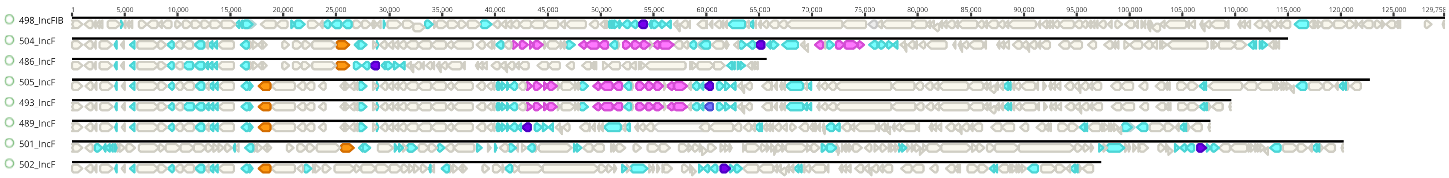


**Figure S4: Overview of IncF plasmids carrying beta-lactamase CTX-M-27**. Transposase and insertion sequences are shown in light blue, antibiotic resistance genes in pink, beta-lactamase CTX-M-27 gene is shown in purple, and plasmid-encoded enterotoxin is colored in orange. Specific antibiotic resistance genes can be found in Table 3. Sequences are not aligned by similarity but organized in the same order as the phylogenetic tree of Figure 1.

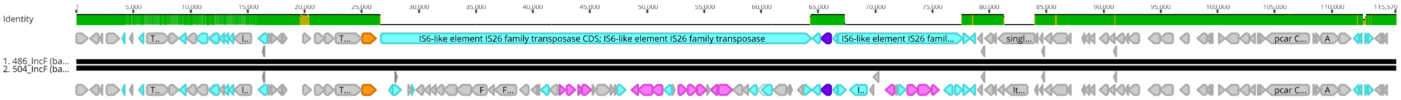


**Figure S5: Alignment of IncF plasmid in isolates 486 and 504**. Alignment region showing insertion sequences (light blue) lacking antibiotic resistance genes that are present in 504 (pink). Beta-lactamase $bla_{CTX\text{-}M\text{-}27}$ gene is shown in purple, also surrounded by IS6 sequences.

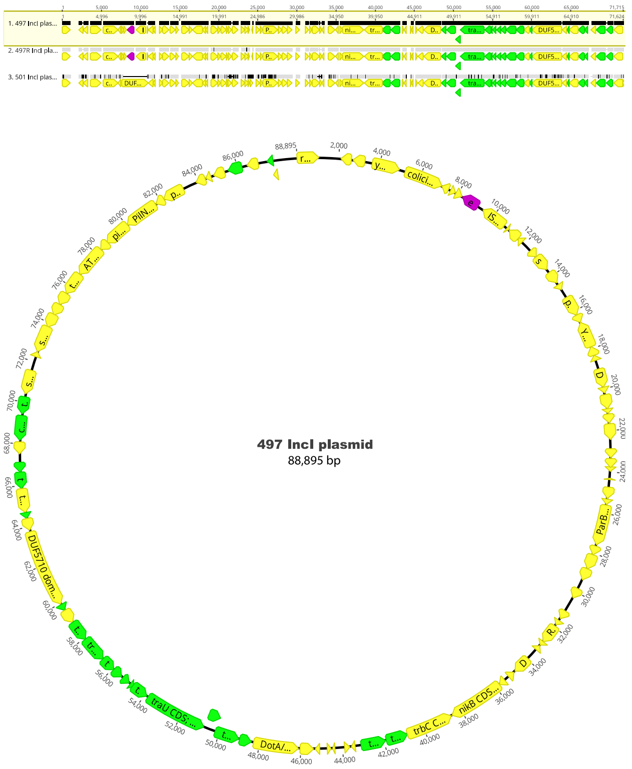


**Figure S6: Map and alignment of plasmid from the IncI1 group shared between different *E. coli* strains of the same participant.** The plasmid replicon of IncI1 was identified by PlasmidFinder. Conjugation-related genes encoded in the plasmid are highlighted in green. Extended-spectrum beta-lactamase gene bla$_{\text{CTX-M-1}}$ is shown in purple. Above, there is an alignment between all IncI1 plasmids found in this study, including plasmids encoded in different *E. coli* isolates from the same participant (497 and 497R).

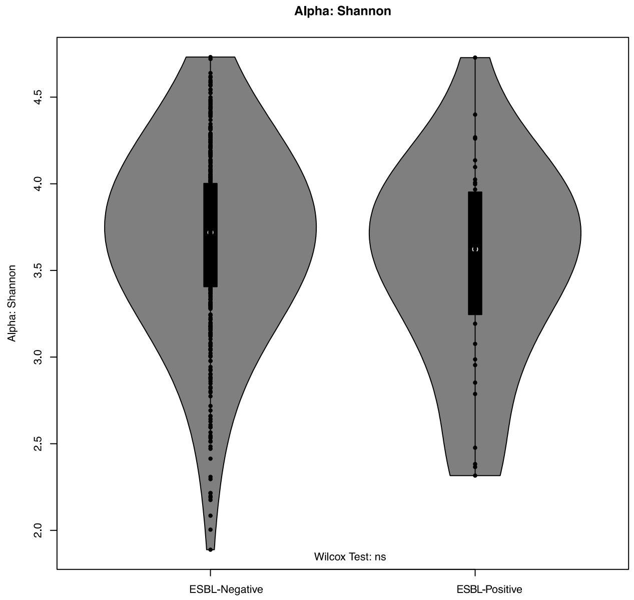


**Figure S7: Alpha diversity by Shannon index.** ASVs found in the initial samples of each participant (555) were tested for alpha diversity comparing ESBL positive and ESBL negative communities. The test was performed using Shannon index in the vegan package of R and confirmed with Wilcoxon test.

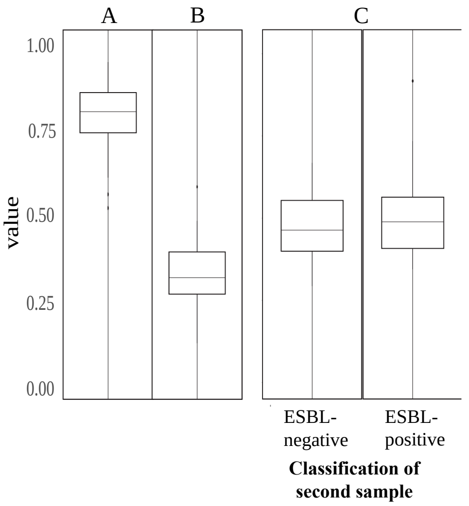


**Figure S8: Bray-Curtis distances between ESBL-positive samples.** Dissimilarity distances calculated between initial samples from each ESBL-positive individual (A), between initial samples and their duplicates (B) and, between initial and second sample from the same individual, grouped by ESBL status of second sample.

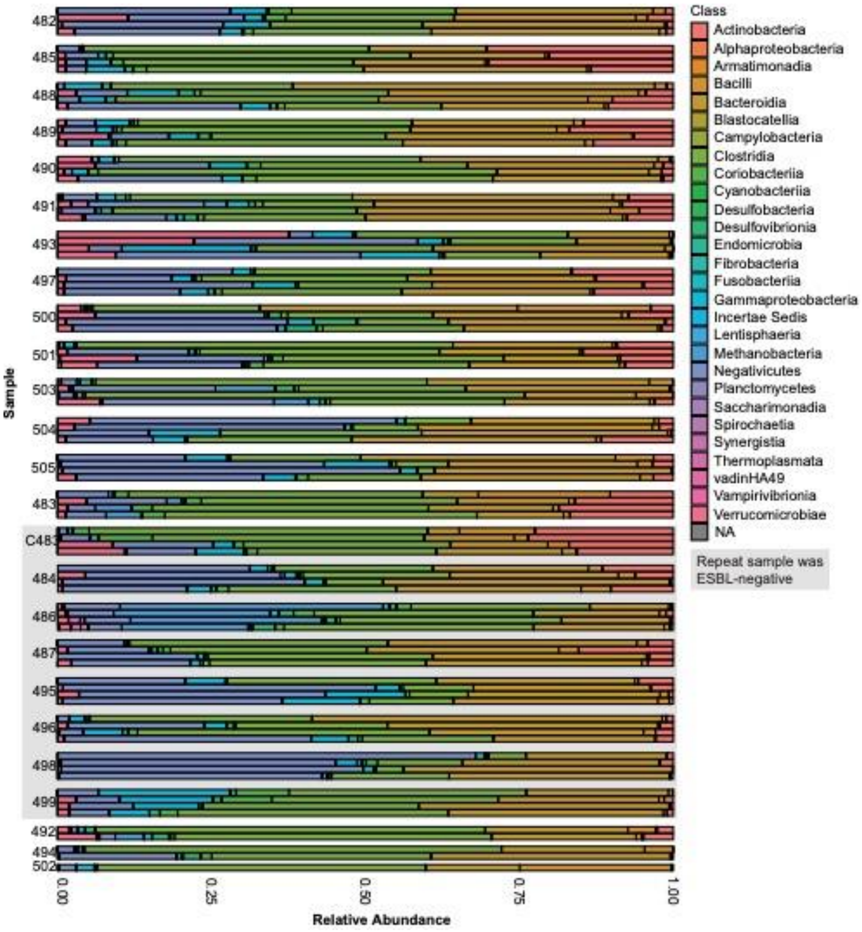


**Figure S9: Microbiota composition of ESBL-positive individuals.** Taxonomic bar plot displaying the relative abundance of Classes in ESBL-positive samples. The number assigned to each cluster represents an individual that was ESBL-positive in their first sample. Each bar cluster includes (from top to bottom): original sample, duplicate of original, repeated sample and duplicate of repeat sample when available. Clusters on the gray box represent individuals that were ESBL-negative on their repeated sample.
